# A multi-ethnic polygenic risk score is associated with hypertension prevalence and progression throughout adulthood

**DOI:** 10.1101/2021.10.31.21265717

**Authors:** Nuzulul Kurniansyah, Matthew O Goodman, Tanika Kelly, Tali Elfassi, Kerri L Wiggins, Joshua C Bis, Xiuqing Guo, Walter Palmas, Kent D Taylor, Henry J Lin, Jeffrey Haessler, Yan Gao, Daichi Shimbo, Jennifer A Smith, Bing Yu, Elena Feofanova, Roelof Smit, Zhe Wang, Shih-Jen Hwang, Simin Liu, Sylvia Wassertheil-Smoller, JoAnn E Manson, Donald M Lloyd-Jones, Stephen S Rich, Ruth JF Loos, Susan Redline, Adolfo Correa, Charles Kooperberg, Myriam Fornage, Robert C Kaplan, Bruce M Psaty, Jerome I Rotter, Donna K Arnett, Alanna C Morrison, Nora Franceschini, Daniel Levy, the NHLBI Trans-Omics in Precision Medicine (TOPMed) Consortium, Tamar Sofer

## Abstract

**Background:** We used summary statistics from previously-published GWAS of systolic and diastolic BP and of hypertension to construct Polygenic Risk Scores (PRS) to predict hypertension across diverse populations.

**Methods:** We used 10,314 participants of diverse ancestry from BioMe to train trait-specific PRS. We implemented a novel approach to select one of multiple potential PRS based on the same GWAS, by optimizing the coefficient of variation across estimated PRS effect sizes in independent subsets of the training dataset. We combined the 3 selected trait-specific PRS as their unweighted sum, called “PRSsum”. We evaluated PRS associations in an independent dataset of 39,035 individuals from eight cohort studies, to select the final, multi-ethnic, HTN-PRS. We estimated its association with prevalent and incident hypertension 4-6 years later. We studied hypertension development within HTN-PRS strata in a longitudinal, six-visit, longitudinal dataset of 3,087 self-identified Black and White participants from the CARDIA study. Finally, we evaluated the HTN-PRS association with clinical outcomes in 40,201 individuals from the MGB Biobank.

**Results:** Compared to other race/ethnic backgrounds, African-Americans had higher average values of the HTN-PRS. The HTN-PRS was associated with prevalent hypertension (OR=2.10, 95% CI [1.99, 2.21], per one standard deviation (SD) of the PRS) across all participants, and in each race/ethnic background, with heterogeneity by background (p-value < 1.0×10^-4^). The lowest estimated effect size was in African Americans (OR=1.53, 95% CI [1.38, 1.69]). The HTN-PRS was associated with new onset hypertension among individuals with normal (respectively, elevated) BP at baseline: OR=1.71, 95% CI [1.55, 1.91] (OR=1.48, 95% CI [1.27, 1.71]). Association was further observed in age-stratified analysis. In CARDIA, Black participants with high HTN-PRS percentiles developed hypertension earlier than White participants with high HTN-PRS percentiles. The HTN-PRS was significantly associated with increased risk of coronary artery disease (OR=1.12), ischemic stroke (OR=1.15), type 2 diabetes (OR=1.19), and chronic kidney disease (OR=1.12), in the MGB Biobank.

**Conclusions:** The multi-ethnic HTN-PRS is associated with both prevalent and incident hypertension at 4-6 years of follow up across adulthood and is associated with clinical outcomes.

## Introduction

Hypertension affects over 1.1 billion people in the world (1). Globally, the number of people with hypertension has increased over time, reflecting the aging of the population and is predicted to reach 1.56 billion people by 2025 (2). Hypertension is a leading risk factor for cardiovascular, kidney, cerebrovascular disease, and a leading cause of global mortality (3–5). The causes of hypertension are genetic and environmental, including dietary factors, and the rising prevalence of obesity (6–8). Genome-wide association studies (GWAS) have identified more than 900 genomic regions associated with blood pressure (BP) phenotypes (9–14), and GWAS from diverse race/ethnic backgrounds as well as admixture mapping studies demonstrate that BP phenotypes have some ancestry-specific or ancestry-enriched genetic components (e.g. genetic variants that are more common in one continental genetic ancestry) (15–20).

Polygenic Risk Scores (PRS) estimate the effect of many genetic variants on an individual’s genetic susceptibility to a disease or trait, typically calculated as a weighted sum of trait-associated alleles, with weights often being the effect estimates corresponding to each allele. PRS are typically constructed using results from GWAS to guide the selection of single nucleotide polymorphisms (SNPs) into the PRS, and their weights (21, 22). Developing PRS that are useful across a diverse, globally representative population remains a challenge when the underlying GWAS is performed primarily in people of European ancestry (23–25). A recent study of PRS for hypertension found that a BP PRS was useful in predicting longitudinal development of hypertension in a Finish population (26). With the availability of recent, large multi-ethnic and non-European GWAS of BP phenotypes, such as from the Million Veteran Program (MVP), the UK Biobank (UKBB), and Biobank Japan (BBJ) (10, 27), it is possible to include SNPs that are common in genetic ancestries that are traditionally less represented in GWAS, permitting the construction of multi-ethnic PRS for hypertension risk prediction (28).

Here we leverage a multi-ethnic dataset, with harmonized genotypes and phenotypes, from the Trans-Omics in Precision Medicine Initiative (TOPMed) program (29, 30) to construct and assess PRS for hypertension based on summary statistics from multiple GWAS of hypertension and BP phenotypes. Individuals were from a few U.S. race/ethnic backgrounds: African Americans (AA), Hispanic/Latino Americans (HA), Asian Americans (AsA), and European Americans (EA), allowing for assessment of the PRS across major U.S. demographic segments. We used multiple independent subsets of the TOPMed dataset to train, test, and assess PRS associations with hypertension across the lifespan. We evaluated the association of the final HTN-PRS with incident outcomes in the Mass General Brigham (MGB) Biobank. To develop the PRS, we propose a new approach for selecting tuning parameters for PRS construction, based on optimizing the coefficient of variation of the effect size estimates of five independent subsets of the training dataset, as well as combination of PRS based on GWAS of multiple BP phenotypes into a single PRS.

## Methods

Figure 1 provides an overview of the study. At stage 1, we used summary statistics from multiple GWAS of BP phenotypes to construct PRS in a training dataset (stage 1 dataset) with prevalent hypertension. We selected GWAS that were based on individuals not overlapping with our dataset (published Million Veterans Program GWAS (10), and summary statistics from FinnGen and UK Biobank databases). We used a clump & threshold methodology which requires selection of tuning parameters. Importantly, we evaluated a few methods to select tuning parameters, and an approach to combine PRS across phenotypes. At stage 2, we further assessed the methods for tuning parameter selection and the combined PRS in a stage 2 dataset using prevalent hypertension at a baseline exam. We selected the best performing PRS, and used it in analyses of PRS association with hypertension using data from two visits in the stage 2 dataset. At stage 3, we studied the PRS association with incident hypertension in young African and European adults, using a longitudinal, stage 3 dataset with 6 exams. At stage 4, we tested the association of the PRS with disease status in individuals from the MGB Biobank (stage 4 dataset).

**Figure 1:**
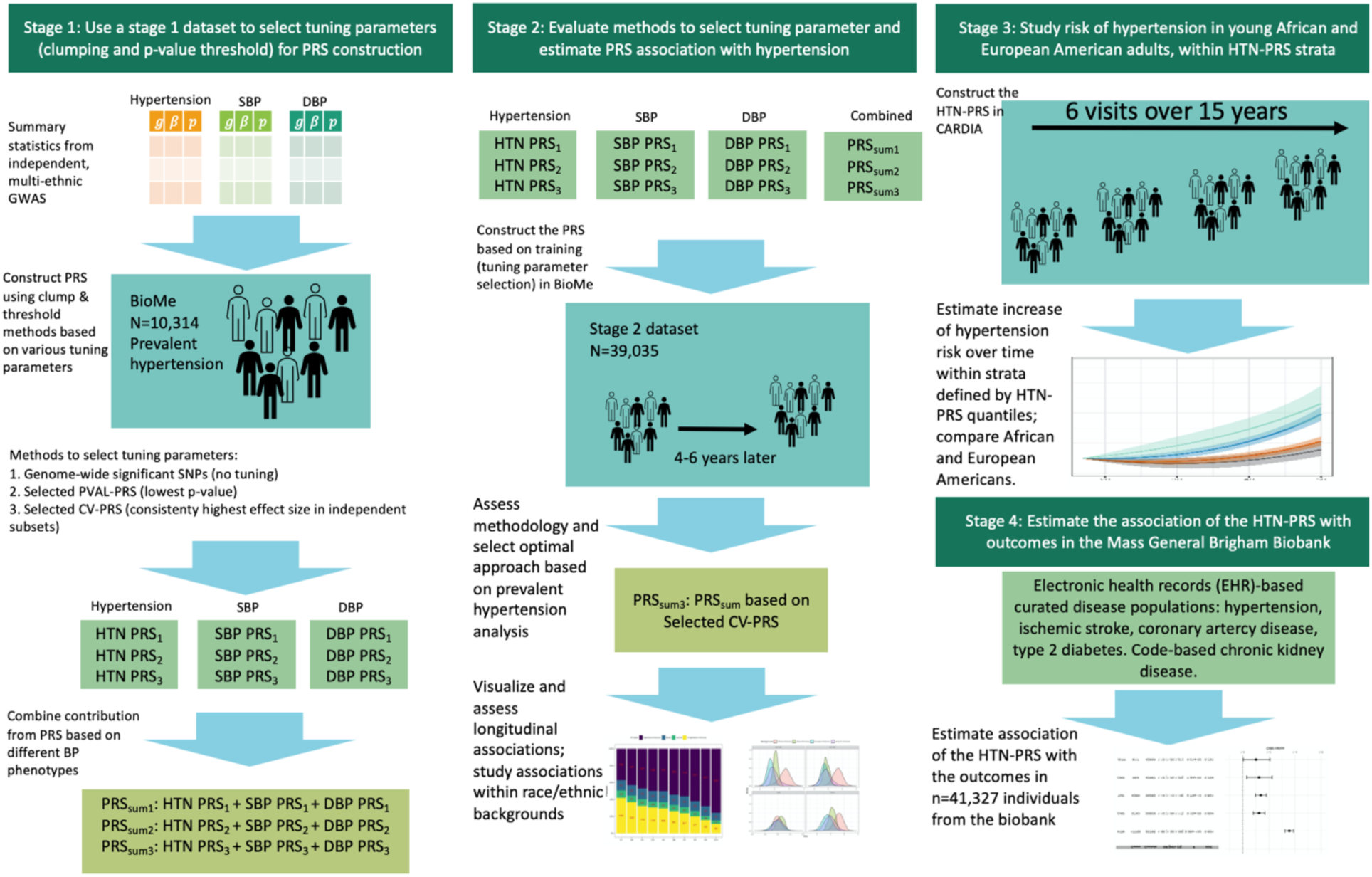
Organization of the study. In stage 1, we used the stage 1 dataset to select tuning parameters for PRS construction based on GWAS of BP phenotypes. We compared a few methods for tuning parameter selection and constructed PRSsum combining a few phenotype-specific PRS. In stage 2 we evaluate the methods for tuning parameter selection in the stage 2 dataset, and selected one PRS, namely HTN-PRS, to move forward for two-visit longitudinal analysis. In stage 3, we used a longitudinal dataset from CARDIA to study hypertension development in young adults, and compared AAs and EAs. In stage 4, we tested the association of the HTN-PRS with disease outcomes in MGB biobank (stage 4 dataset).

### The TOPMed Dataset

The TOPMed dataset included 52,436 individuals from 10 TOPMed studies. Based on the characteristics of these studies, the TOPMed data was split into stage 1, stage 2, and stage 3 datasets. Stage 1 dataset was the Mount Sinai BioMe Biobank, which included 10,314 diverse individuals with prevalent hypertension status. It was used as a training dataset for constructing the hypertension PRS. Stage 2 dataset included 39,035 individuals from an additional eight studies, with all individuals being genetically unrelated (at the third degree) to those in the training dataset. Stage 2 dataset was longitudinal, with hypertension status assessed in two exams, on average 4-6 years apart. Individuals were self-reported from four predominant U.S. race/ethnic backgrounds, with 22,701 EA, 8,822 AA, 6,718 HA, and 794 AsA individuals. Stage 3 dataset included the CARDIA study, with 6 exams over 15 years follow up of young Black and White individuals, and was used to compare the development of hypertension risk within PRS strata across the two race/ethnic backgrounds.

### Prevalent and longitudinal measures of hypertension

Systolic BP (SBP) and diastolic BP (DBP) were measured in each study according to methods provided in the study descriptions in the Supplementary Materials. Hypertension stages were defined according to (1) Normal BP: SBP ≤ 120 mmHg and DBP ≤ 80 mmHg and untreated; (2) Elevated BP: SBP between 120-129 and DBP ≤ 80 mmHg, and untreated; (3) Hypertension: SBP ≥ 130 mmHg, DBP ≥ 80 mmHg, self-reported physician diagnosed hypertension, or use of anti-hypertensive medications (31). When examining 2-exams longitudinal patterns of hypertension using the stage 2 dataset, we performed data visualization in which we categorized individuals as: not having hypertension across the two exams (healthy longitudinal trajectory; may include individuals with normal and with elevated BP, but without change in these categories between the exams); having hypertension in the two exams (severe longitudinal trajectory); BP category worsen between exams, including individuals who had normal BP at the baseline exam, and elevated BP or hypertension at the follow-up exam, or elevated BP followed by hypertension; BP category improved between exams, including individuals who were not treated by antihypertensive medications in any of the exams, and had improved BP category (hypertensive to elevated or normal, or elevated to normal). Individuals treated with antihypertensive medication in either baseline or follow-up exam were never categorized as “improved”. We also performed association analysis of the PRS with new onset hypertension at the follow-up exam, focusing, separately, on individuals who had normal BP at baseline and who had elevated BP at baseline.

### Whole Genome Sequencing

We used whole genome sequencing data from the Trans-Omics in Precision Medicine (TOPMed) program Freeze 8 release (29). Only variants with minor allele frequency (MAF) ≥ 0.01 were used in this analysis. Information about genome sequencing, variant calling, and quality control procedures is available here https://www.nhlbiwgs.org/topmed-whole-genome-sequencing-methods-freeze-8. The TOPMed Data Coordinating Center constructed a sparse kinship matrix estimating recent genetic relatedness where values were set to zero when the genetic relationship was estimated to be more distant than 4^th^degree relatedness, and principal components (PCs), using the PC-Relate algorithm (32).

### Published GWAS of BP Phenotype

Table 1 provides information about hypertension and BP GWAS used to construct PRS. In primary analysis, we used multi-ethnic GWAS: hypertension “pan ancestry” GWAS from UKBB (https://pan.ukbb.broadinstitute.org/), and systolic BP (SBP), and diastolic BP (DBP) from MVP (10). Note that UKBB pan ancestry GWAS are multi-ethnic, however, U.S. minorities are not well represented compared to MVP, and therefore we prioritized MVP as a primary GWAS for SBP and DBP. All these GWAS are based on large sample sizes and have no overlap in participants with the TOPMed BP dataset. In secondary analysis, we also used hypertension GWAS from FinnGen (https://www.finngen.fi/en) and SBP and DBP GWAS from UKBB pan ancestry and BBJ (33), and performed inverse-variance fixed-effects meta-analyses using METAL (34) for each BP trait GWAS (SBP, DBP and hypertension). These are described in Supplementary Table S1. Secondary analyses were only performed on the training dataset.

**Table1.** External GWAS used for hypertension PRS construction

| GWAS name | Reference | Trait | Sample Size | Population |
| --- | --- | --- | --- | --- |
| MVP | PMID:30578418 (10) | SBP | 318,492 | Multi-ethnic (69.1% non-Hispanic Whites, 18.8% non-Hispanic Blacks, 6.7% Hispanics, 0.77% non-Hispanic Asians and 0.85% non-Hispanic Native Americans) |
|  |  | DBP | 318,891 |  |
| Pan UKBB | No manuscript, downloaded from https://pan.ukbb.broadinstitute.org | HTN | 451,894<br>(126,196 cases, 325,698 controls) | African (1.43 %), Admix American (0.21 %), European (86.75 %), Central/South Asian (1.89 %), Middle Eastern (0.34%), East Asian (0.57%) |
|  |  | SBP | 453,005 |  |
|  |  | DBP | 453,010 |  |

The table provides GWAS source, study population as reported by the manuscript or repository reporting the GWAS, and number of participants used to generate summary statistics.

### Quality control on summary statistics

We filtered SNPs with MAF<0.01 in the discovery GWAS from Table 1 and/or in the dataset comprising of all TOPMed analysis participants (stage 1, 2 and 3 datasets combined), and SNPs that did not pass TOPMed quality filters. We re-coded the variant positions and alleles to match those in the TOPMed data (via the UCSC hg19 to hg38 chain file) using rtracklayer R package version 1.46.0 (35)

### PRS construction based on a single GWAS

We constructed PRS using the clump-and-threshold method implemented in the PRSice 2 software version v2.3.3 (22) using each of the GWAS in Table 1. Three tuning parameters are required: p-value threshold, and two clumping parameters. As p-value thresholds, we used 5×10^-8^, 1×10^-7^, 1×10^-6^, 1×10^-5^, 1×10^-4^, 1×10^-3^, 1×10^-2^, 0.1, 0.2, 0.3, 0.4, 0.5. For clumping, we used the entire TOPMed dataset (stage 1, 2, and 3 datasets combined) as a reference panel for Linkage Disequilibrium (LD) and set clumping parameters to R^2^=0.1, 0.2 and 0.3 and distance 250kb, 500kb, and 1000Kb. To standardize PRS while keeping effect sizes comparable in all analyses, we computed the mean and standard deviations (SDs) of each type of PRS based on the complete TOPMed dataset. Then, we used these means and SDs in all subsequent analyses: for a given PRS in any dataset, we subtracted its pre-computed mean and divided it by its pre-computed SD. This standardization approach allowed for obtaining comparable effect sizes across stage 1, 2, 3, and 4 datasets, as well as across background-specific and multi-ethnic analyses. Standardization does not influence p-values or prediction measures.

### Using stage 1 dataset to select of tuning parameters for PRS construction

To select tuning parameters for a PRS based on a given GWAS from Table 1 (or based on meta-analysis of multiple GWAS), we examined the association of various constructed PRS with prevalent hypertension in the stage 1 dataset. We developed the selected CV-PRS approach, which we describe below, and compare it to two additional widely-used approaches: genome-wide significance PRS, selected PVAL-PRS. Both the selected CV-PRS and the selected PVAL-PRS approaches attempt to select one set of tuning parameters (p-value threshold and LD clumping parameters) to construct PRS out of all tuning parameter combination used. The selected CV-PRS approach aims to identify the tuning parameters that yield consistently high PRS effect size in new, independent, datasets. To do this, it minimized the coefficient of variation (CV) computed on the PRS effect size estimates obtained from 5 equal-sized independent subsets of BioMe (this is visualized in Figure 1). Specifically, the CV was estimated as the standard deviation of the five effect (log odds ratio) estimates, divided by the mean of these effect estimates. The selected PVAL-PRS is the PRS with the lowest association p-value in the stage 1 dataset. The genome-wide significance PRS was constructed using SNPs with p-value<5×10^-8^, and fixed clumping parameters: R^2^=0.1 and distance of 1000kb, and otherwise no selection of other parameters.

### PRS construction based on multiple GWAS

We constructed a PRS called “PRSsum” by summing PRS constructed based on the three BP phenotypes (SBP, DBP, hypertension). The three PRS were summed after each was scaled using the mean and SD computed using the entire TOPMed dataset. We summed non-adaptively, i.e., unweighted sum with PRSsum = PRS1 + PRS2 + PRS3. We generated three PRSsum, based on the three potential strategies to construct PRS: 1) PRSsum based on genome-wide significance (which summed genome-wide significant PRS); 2) PRSsum based on PVAL-PRS (which summed the selected PVAL-PRS across the three phenotypes), and 3) PRSsum based on selected CV-PRS (which summed the CV-PRS from the three phenotypes). The final multi-ethnic HTN-PRS was the one based on the approach that consistently performs better on the test dataset with prevalent hypertension. We used it for follow up analysis of longitudinal measures of hypertension, and in stage 3 and 4 analyses (Figure 1)

### Association analysis of PRS with hypertension in stage 1 and 2 datasets

We used logistic mixed models, as implemented in the GENESIS R package (36) version 2.16.1 to estimate the association between the PRS and hypertension, with relatedness modeled via a sparse kinship matrix. Association analyses were adjusted for age, age^2^, BMI, smoking status (current smoker versus former or never smoker) at the baseline exam, the first 11 PCs, race/ethnicity when evaluating PRS association in a multi-ethnic sample, and time between exams when studying incident associations. We performed two analyses of incident, new onset hypertension in the follow-up exam: one based on individuals who had normal BP at baseline, and second based on individuals who had elevated BP at baseline. We accounted for differences in variances across groups defined according to self-reported race/ethnicity and study by estimating a different residual variance parameter for each (37). We estimated both multi-ethnic and background-specific PRS associations. For the latter, we tested for heterogeneity of estimated effects by race/ethnic background using the Cochran Q test that accounts for covariance between effect estimates across the background groups (38). We evaluated the predictive performance of PRS by calculating the area under the receiver operating curve (AUC) using the AUC function from the pROC R package (39), version 1.16.2. We used only unrelated individual when calculating AUC. We visualized the unadjusted association of the final HTN-PRS with longitudinal measure of hypertension via a decile plot, demonstrating proportions of individuals in categories of longitudinal BP trajectories in each of the PRS deciles. In secondary analysis, to benchmark the effect of the HTN-PRS against known hypertension risk factors, we compared standardized effect size estimates of the HTN-PRS, BMI, and smoking status, from both the prevalent and incident hypertension analyses.

### Development of new onset hypertension in young adulthood by PRS levels

Stage 3 dataset consisted of n=1,388 self-identified Black and n=1,699 self-identified White young adults from CARDIA. Follow up started on average at age 25 (minimum age of participants at baseline = 17). We used 15 years of follow up available on the dbGaP repository (40). We generated the HTN-PRS for each of CARDIA participant, removed related individuals (degree 3 or higher) and assigned individuals to strata defined by <10 percentile of the PRS, 10-50, 50-90, and >90 percentiles. We first computed these strata across all CARDIA individuals. Next, because there was only a single Black individual in the bottom stratum and only a single White individual in the top stratum, we also defined strata within Black and White groups separately. Next, we fit generalized linear mixed models (GLMM) with random intercept for each participant within strata in the combined and background-specific analyses. The outcome was hypertension, defined as before, and the exposure variables were sex, 11 PCs, age, and age squared (PRS values were not included as explanatory variables). We subtracted the minimum age in the sample, 17, from all age values, for ease of computation of effect later on. The effect estimates of age and age squared are denoted by 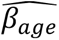 and 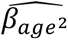. We used the model to estimate the odds ratio (OR) for hypertension by age relative to the minimum age in the sample, based on the coefficients from the GLMM, i.e. [(*age* − 17) × 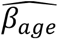 + (*age* − 17)^$^ × 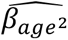] and computed a 95% confidence interval separately for each age, by computing standard errors (SEs) for the above equation based on the estimated SEs and covariances of 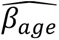 and 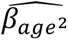.

### Association of the HTN-PRS with outcomes in the MGB Biobank

We tested the association of the HTN-PRS with hypertension (another form of replication), coronary artery disease (CAD), ischemic stroke, type 2 diabetes, and chronic kidney disease (CKD), in the MGB Biobank (stage 4 dataset). We also tested the association of the HTN-PRS with obesity because the pan-UKBB GWAS summary statistics that we used for constructing one of the PRS used by the HTN-PRS was from an analysis not adjusted to BMI. We used n=40,201 unrelated individuals with relevant phenotypes. For all outcomes other than CKD we used “curated disease populations” defined by a phenotyping algorithm that uses ICD-9 codes and natural language processing (41). For CKD we used a single term referring to having a health care system encounter due to CKD (“reason to visit” is CKD). We used logistic regression adjusted for 10 PCs, current age, sex, race/ethnicity. The main analysis was multi-ethnic, and we also tested race/ethnic background-specific associations, though sample sizes were small in non-EA groups. Comprehensive description of the MGB Biobank methods is provided in the Supplementary Information.

## Results

Table S2 characterizes the stage 1 dataset, used for training the PRS using prevalent hypertension analysis. Rates of hypertension among the race/ethnic groups ranged from 56% with 32% treated (AsA) to 79% with 57% treated (AA). Mean age ranged from 53 (AsA) to 58 (HA). Table S3 characterizes the sample across the eight studies participating in the stage 2 dataset. The data were collected over two time-points with an average of 4-6 years between measures. There were 39,035 individuals in the analytic sample, of which 22,701 were EA, 8,822 were AA, 6,718 were HA, and 794 were AsA. The characteristics of the race/ethnic background groups were quite heterogeneous. The average age across backgrounds ranged from 51 (HA) to 62 (AsA) at baseline. The EA group had the highest proportion of female participants (72.2%) while the HA group had the lowest (62%). The number of hypertension cases increased between the exams in all background groups. AAs had the highest proportion of hypertension cases in both exams: 76.5% and 53% treated (baseline), and 83.3% and 66% treated (follow-up). HAs had the fewest cases: with 51.7% and 23% treated (baseline) and 59.6% and 39.5% treated (follow-up).

### PRS tuning parameters selection based on stage 1 dataset

Based on each GWAS, we selected PRS using three criteria: Genome-wide Significant PRS; Selected CV-PRS; Selected PVAL-PRS. Supplementary Figure S1 describes the association of each PRS with hypertension in the stage 1 dataset. Selected CV-PRS had the highest AUC compared to other PRS. Supplementary Figure S2 further reports these associations for the secondary GWAS as well, showing that they mostly performed less well than the primary GWAS, with the exception of the PRS based on the UKBB+ICBP EA GWAS (9), in which all TOPMed EA individuals participated. The meta-analysis of all available independent GWAS performed less well than the primary GWAS. Table S4 reports the clumping parameters and number of SNPs for each of the primary and secondary GWAS and each selection criterion.

### PRS associations with baseline hypertension in the stage 2 dataset

Figure 3 demonstrated the trained PRS associations with prevalent hypertension at baseline in the stage 2 dataset. PRS were associated with prevalent hypertension and showed a similar association patterns as in the training dataset, with the exception that here Selected CV-PRS often having higher OR and AUC for each PRS. This pattern was more pronounced for SBP and PRSsum. Here, Selected CV-PRS often had lower p-value compared to the Selected PVAL-PRS. PRSsum based on selected CV-PRS had the strongest association with hypertension (OR=2.10, 95% CI [1.99, 2.21], p-value <1×10^-100^, AUC=0.76). Based on these results, we move forward with PRSsum based on Selected CV-PRS for analysis of incident hypertension. Figure 4 shows the distributions of Selected CV-PRS based on each GWAS and their combined PRSsum. For all PRS, the AA group tended to have the highest PRS values. We computed the correlation between PRS and stratified by race/ethnic background as described in supplementary S5. As expected, PRSsum based on Selected CV showed a strong correlation with each PRS. In what follows, we refer to PRSsum based on Selected CV-PRS as the HTN-PRS, for brevity.

**Figure 2:**
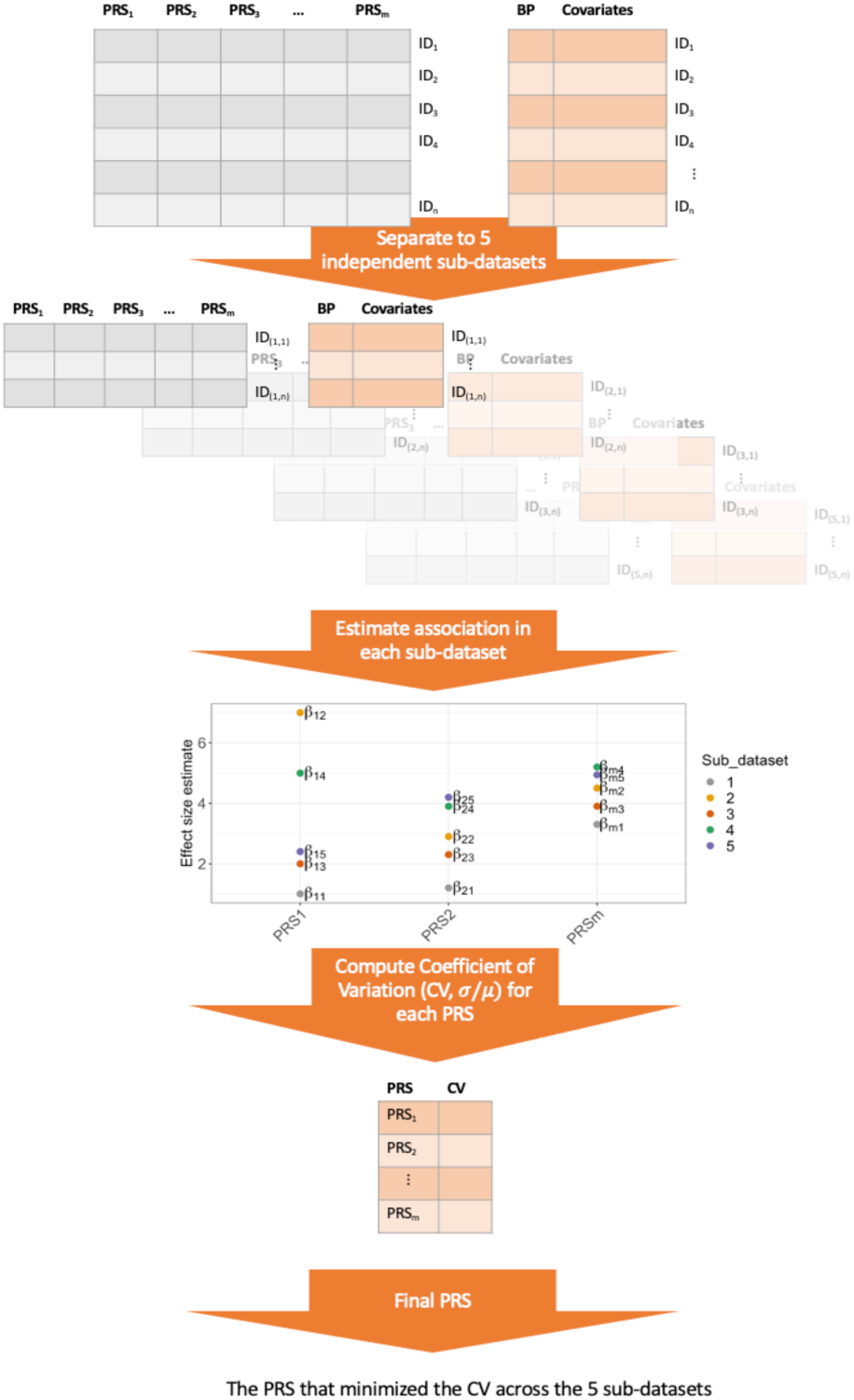
PRS selection using coefficient of variation workflow. Flowchart describing the selection of PRS according to the coefficient of variation (CV) criterion. The data set is split into five independent sub-datasets (without related individuals between the subsets). An association model is fit on each sub-dataset for each PRS. Each PRS, defined by a unique combination of tuning parameter, has 5 independent effect size estimates. We compute the CV for each such PRS, and select the PRS that minizes the CV.

**Figure 3:**
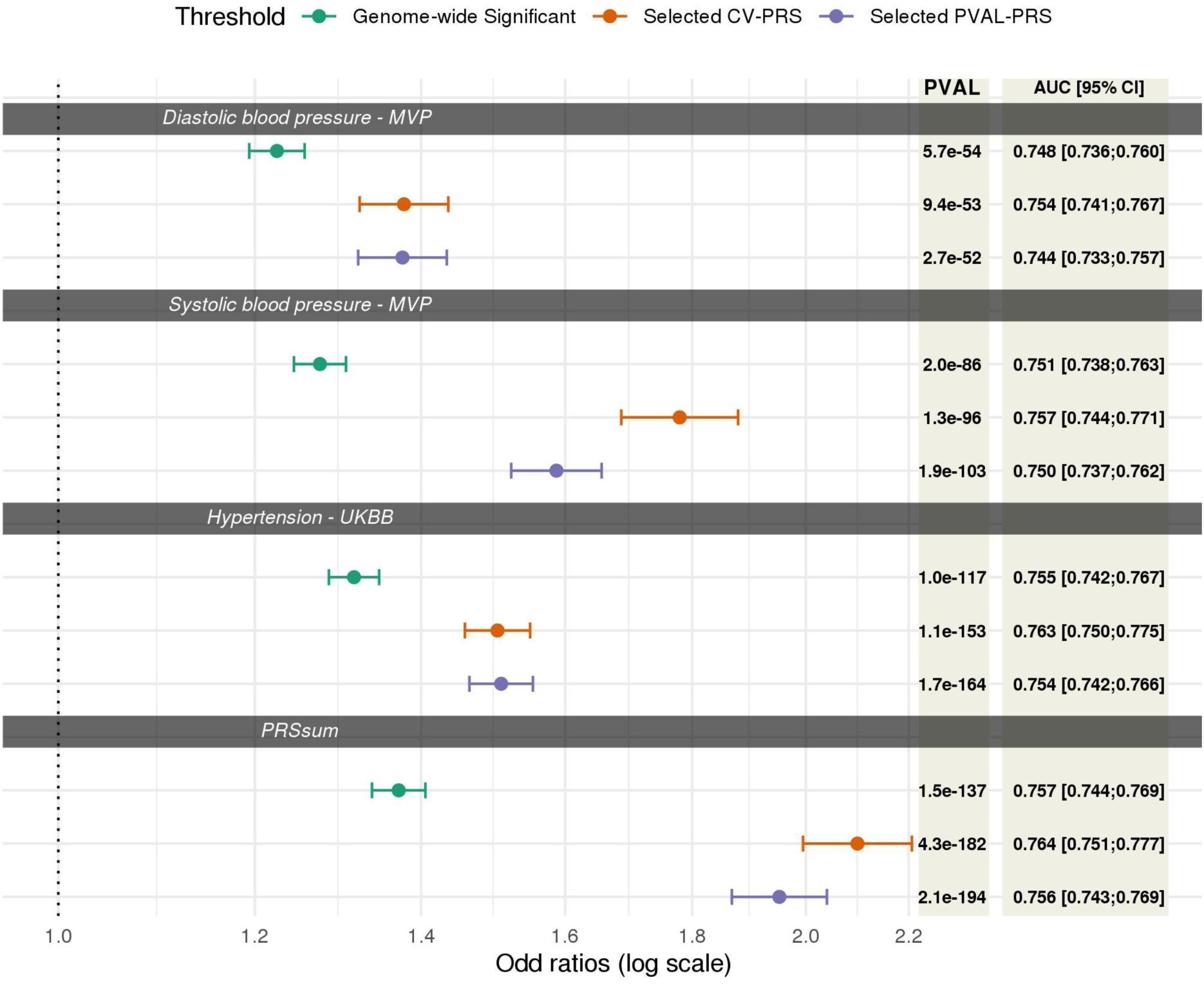
Association of PRS with prevalent hypertension at baseline in the stage 2 data set. Associations of PRS in stage 2 dataset. PRS were trained for hypertension association using stage 1 dataset. “Genome-wide significant PRS” are PRS constructed using genome-wide significant SNPs in the discovery GWAS, with fixed LD parameters or R^2^=0.1 and distance =1000kb. “Selected CV PRS” are PRS that minimized the coefficient of variation (CV) across effect size (log odds ratio (OR)) estimates in 5 independent subsets of the stage 1 dataset. “Selected PVAL-PRS” are PRS that minimized the association p-value with hypertension in the stage 1 dataset. The figure provides estimated ORs, 95% confidence intervals, p-values from association with hypertension, and Area Under the Receiver Operator Curve (AUC). The PRS association was estimated in a model adjusted for sex, age, age^2^, study site, race/ethnic background, smoking status, BMI, and 11 ancestral principal components.

**Figure 4.**
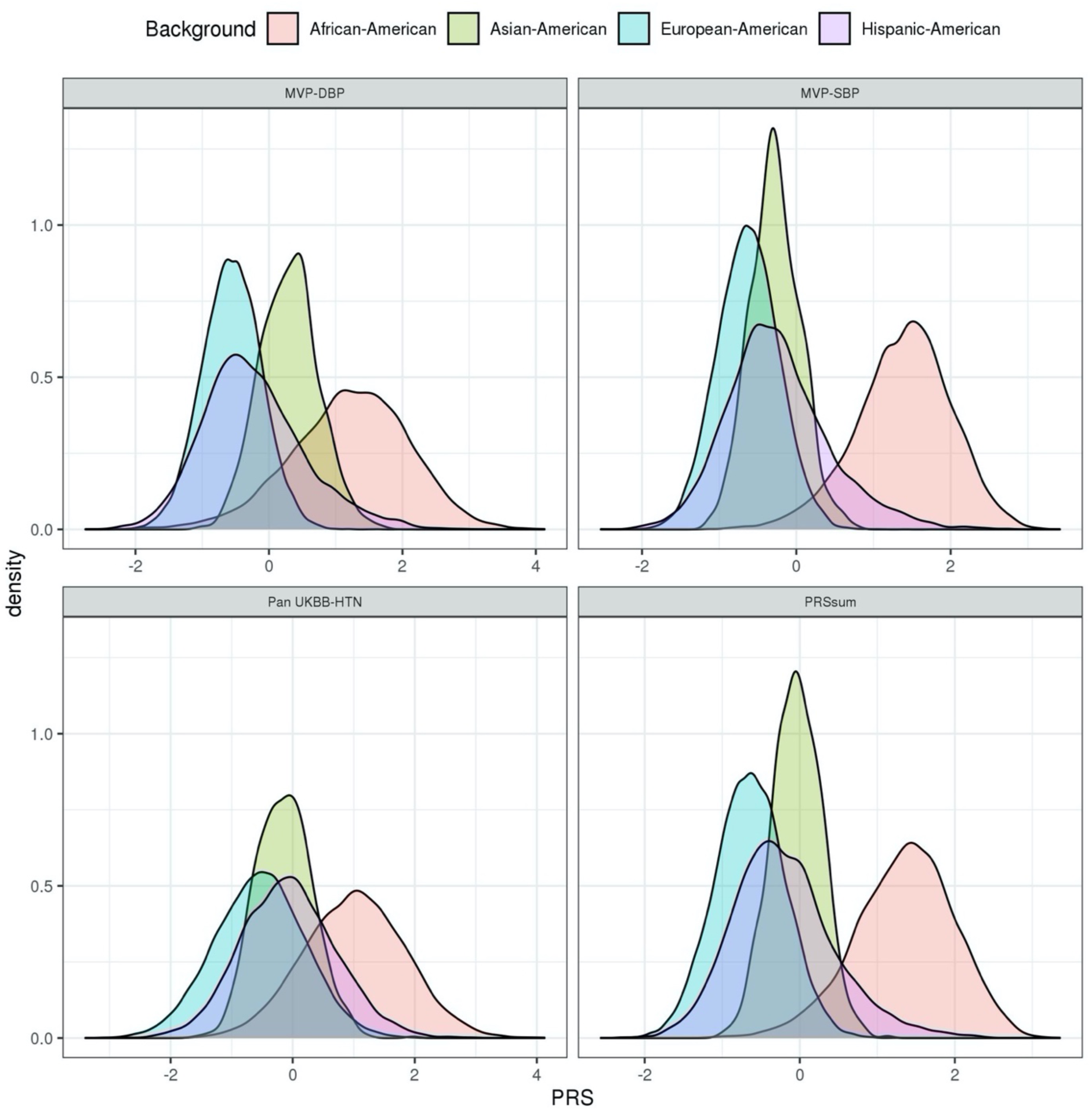
PRS distribution stratified by race/ethnic background. Density plots showing the distributions of Selected CV-PRS based on each GWAS used (Table 1) and PRSsum constructed by summing Selected CV-PRS from the three GWAS (the final HTN-PRS). The figures was created using the stage 2 dataset. The densities are stratified by race/ethnic background.

### Distributions of longitudinal BP categories across deciles of the HNT-PRS

Figure 5 visualizes the distribution of the longitudinal BP categories across deciles of the HTN-PRS. Indeed, higher deciles have higher proportions of individuals with severe BP category (having hypertension already at baseline), and lower deciles have higher proportions of individuals who were free of hypertension in both exams. Relatively few individuals were categorized as “worsened” or “improved” (transitioning between normal BP, elevated BP, and HTN between exams) and there was no strong pattern in their distributions across deciles of the HTN-PRS, with perhaps less individuals in the “improved” category in the higher HTN-PRS deciles compared to the lower ones. Supplementary Figure S3 shows similar data stratified by race/ethnic background and demonstrates generally similar patterns across backgrounds, except for AsA, who are also the group of the smallest sample size (n=794), and therefore there is higher uncertainty in results for this group. Supplementary Figure S4 visualizes similar data stratified by age decades at baseline (≤20, 21-30, 31-40, … 71-80, >80). We observed longitudinal associations of the HTN-PRS with BP category is most age groups (age 31 to age 80), with most the pronounced associations from ages 41-70, for which each severity category is well represented in the data.

**Figure 5:**
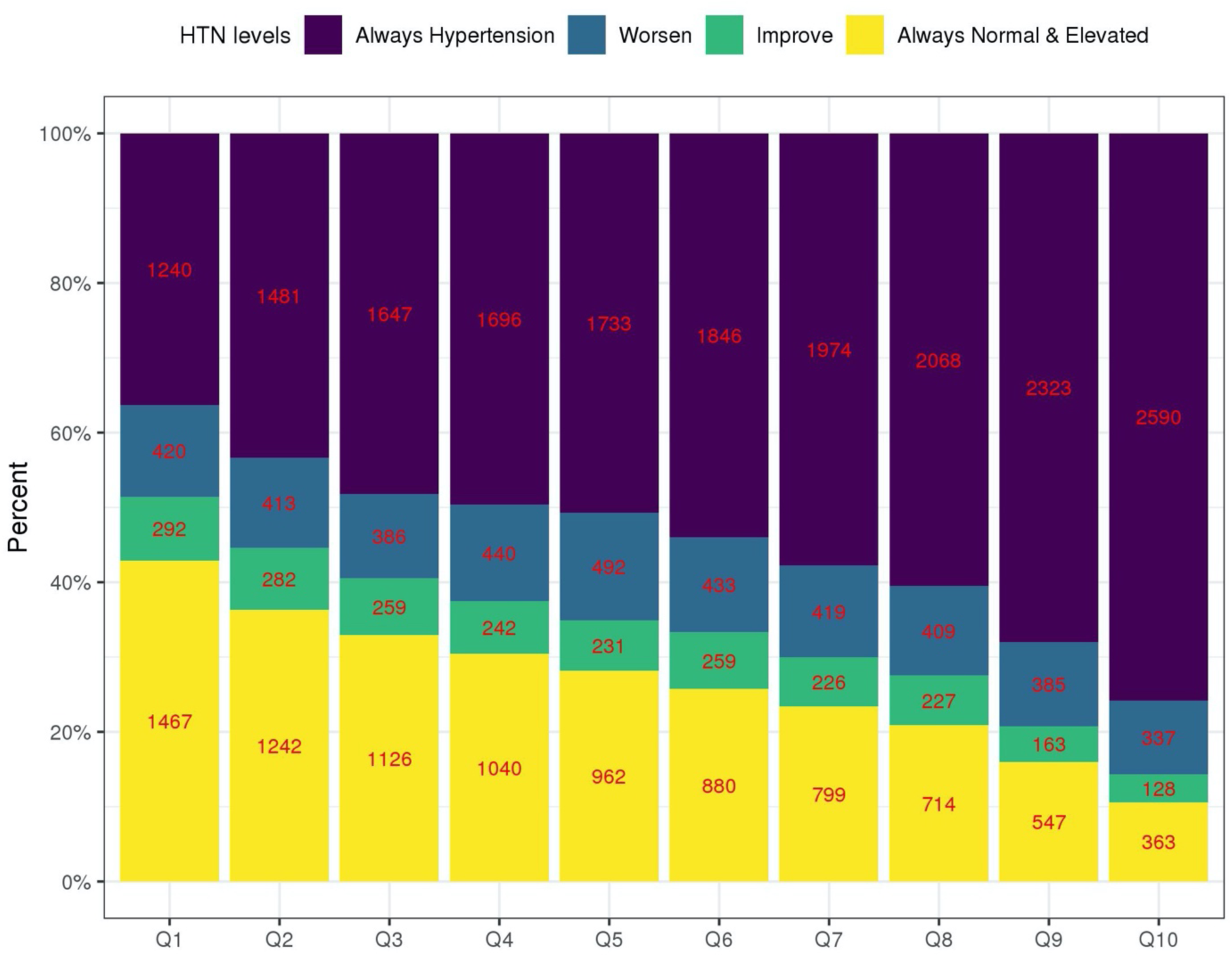
Distribution of longitudinal categories of BP by deciles of the HTN-PRS. The figure visualizes the distribution of longitudinal BP categories in the stage 2 dataset: hypertension at both exams (treated and hypertension in both exams), worsen (individuals having worse BP category in the follow-up exam compared to the first) improved (individuals having better BP category in the second exam compared to the first, only if they were not treated for hypertension at any point), and no hypertension in both exams (includes normal and elevated BP but without change in category), in deciles of the multi-ethnic HTN-PRS. The numbers provide the sample sizes represented by each bar.

### HTN-PRS association with prevalent and incident hypertension across race/ethnic backgrounds

Figure 6 demonstrates the association of the HTN-PRS with three hypertension measures: prevalent hypertension at baseline, new onset hypertension among individuals with normal BP at baseline, and new onset hypertension among individuals with elevated BP at baseline. In the multi-ethnic analysis, the PRS was associated with each of the measures, with strongest association with hypertension at baseline (OR=2.10, 95% CI [1.99, 2.21], p-value <1×10^-100^, AUC=0.76), while new onset hypertension among individuals with normal BP at baseline had OR=1.72, 95% CI [1.55,1.91], p-value=4.67×10^-24^, AUC=0.66, and among those with elevated BP at baseline had OR=1.48, 95% CI [1.27,1.71], p-value=2.39×10^-7^, AUC=0.59). Stratifying the association by race/ethnic background and testing for heterogeneity suggested differences in the PRS association with hypertension at baseline (heterogeneity p-value < 1.0×10^-4^) but weak evidence for heterogeneity otherwise, perhaps due to decreased sample sizes and lower power. Overall, the PRS had the weakest estimated effect sizes, for all outcomes, in the AA group. In secondary analysis, we computed the risk of hypertension at baseline in top versus bottom decile of the PRS within each race/ethnic background. Results are provided in Supplementary Figure S6. The association was strongest in the HA group (OR=4.33, 95% CI [2.81, 6.68]) followed by the EA, AsA, and AA groups.

**Figure 6:**
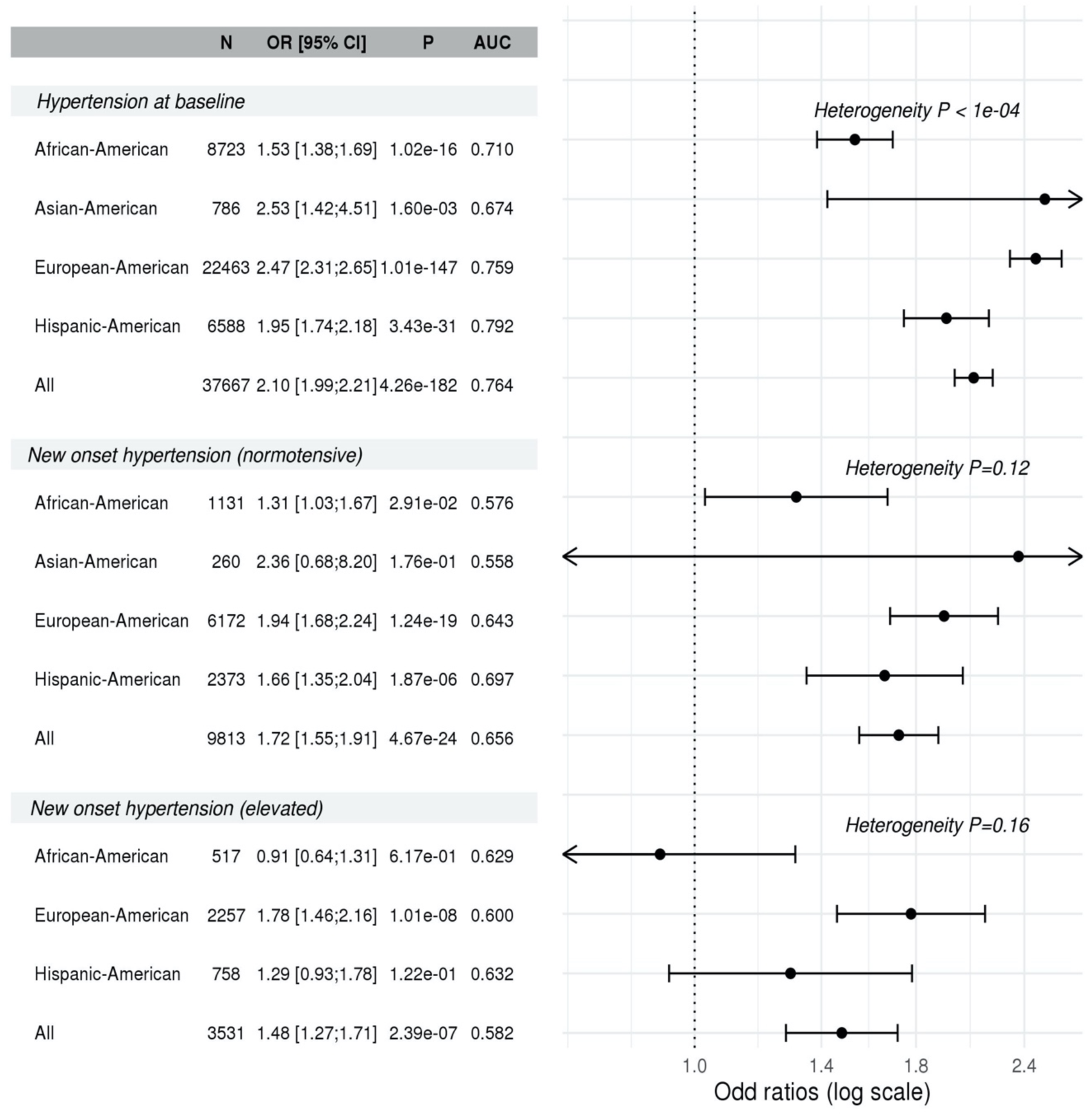
Association of HTN-PRS with hypertension measures across race/ethnicities. The forest plot provides the association of the HTN-PRS with prevalent and incident hypertension in the stage 2 dataset, and within race/ethnic backgrounds. The top part corresponds to prevalence analysis at the baseline visit, the middle part corresponds to prediction of new onset hypertension in exam 2, among individuals who had normal BP at baseline, and the bottom part corresponds to prediction of new onset hypertension in exam 2, among individuals who had elevated BP at baseline. For each analysis we provide sample size, estimated odds ratio (OR) and 95% confidence interval, p-value, and area under the receiver operating curve (AUC). Heterogeneity of effects across race/ethnic groups was tested using the Cochran’s Q test accounting for correlation due to genetic relatedness across groups.

**Figure 7:**
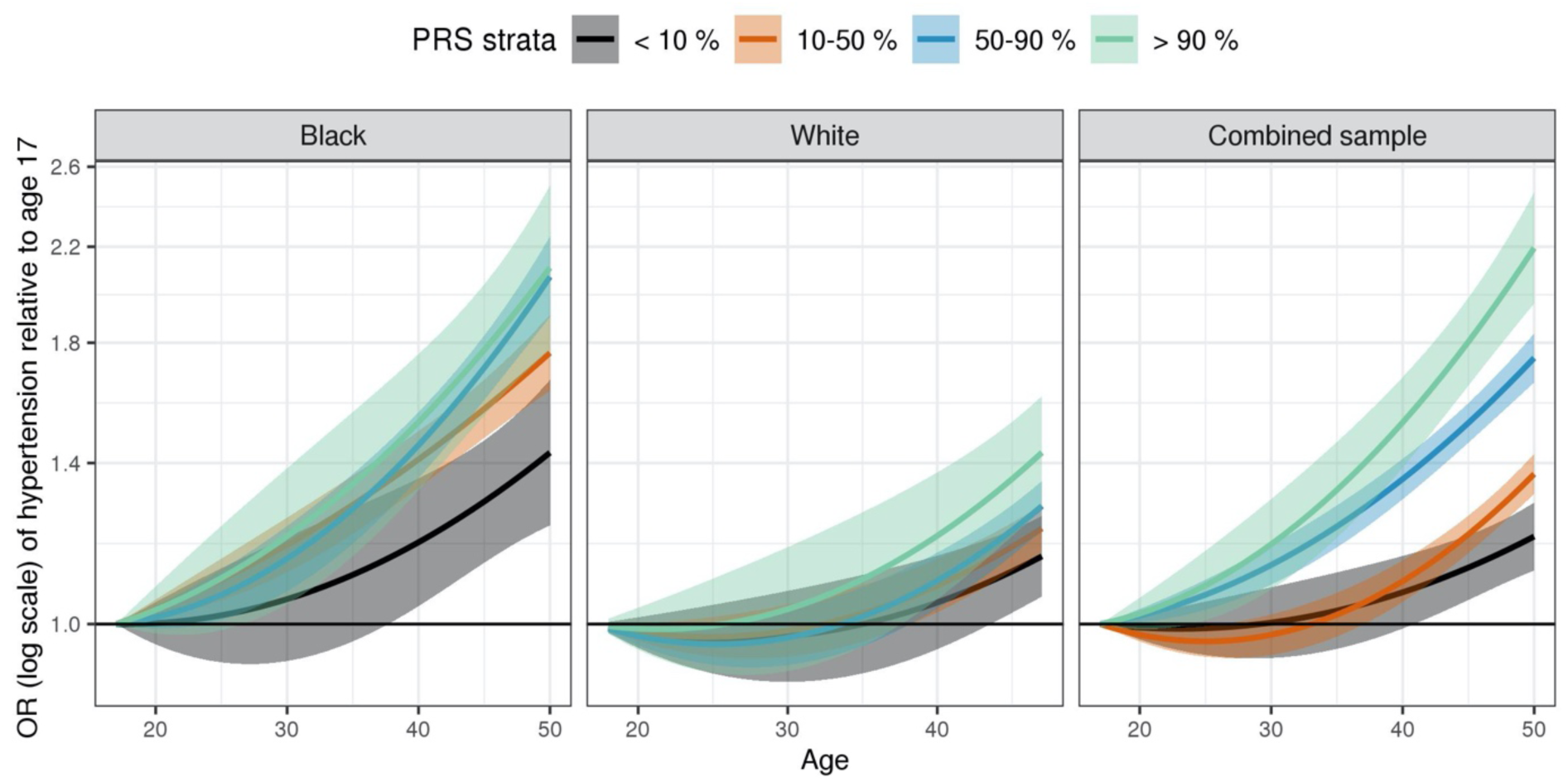
Trajectories of hypertension risk by strata defined by HTN-PRS in young adults from CARDIA. Results from analysis of age-dependent risk of hypertension in young adults from CARDIA (stage 3 dataset). We used generalized linear mixed model to model the risk of hypertension by age within strata defined by quantiles of the HTN-PRS. Analyses were adjusted for age, sex and the first 11 PCs of genetic data. Stratification by PRS quantiles was performed in each presented group: All (combined Black and White participants), Black, and White. The effect of age was modelled using a second degree polynomial. At each point on the curve we provide 95% confidence interval of the effect estimate. In the combined sample, all individuals in the top PRS strata are Black, and 99 % of the individuals in the bottom PRS strata are White.

Supplementary Figure S7 further provides results from association analyses stratified by age decade at baseline. Significant (p<0.05) associations with prevalent and with new onset hypertension are observed throughout adulthood, starting with the 21-30 age group, and up to the 71-80 age group, with the exception that association of incident hypertension among individuals with elevated hypertension in the 21-40 and 51-60 age groups were not statistically significant. This could be due to low sample sizes (see figure for more details).

Supplementary Figure S8 provides a comparison of effect sizes and predictive performance measured by AUC of the multi-ethnic PRS, BMI, and current smoking, in prevalent and incident hypertension analyses. The PRS is comparable to BMI, and both perform better than current smoking.

### PRS Association with development of hypertension in young Black and White adults

We estimated trajectories of hypertension development as second order polynomial functions of age within strata defined by quantiles of the PRS. As shown in Figure 6, in the combined analysis of both Black and White CARDIA participants, trajectories of hypertension risk are separated between the PRS-defined strata, with individuals in strata defined by higher PRS values consistently develop higher hypertension risk compared to those in lower PRS strata. Note that Black individuals obtain higher PRS values compared to White individuals and vice versa. Looking at the race-defined strata, this pattern is seen much more clearly in the Blacks, but less so in Whites alone, suggesting that Blacks with high PRS values (compared to other Blacks) are likely to develop hypertension earlier than Whites with high PRS values (compared to other Whites). At age 50, Black individuals at the 90-100% percentile of the HTN-PRS had 2.11 OR (95% CI [1.77,2.50]) for the risk of hypertension compared that in age 17, individuals at the 50-90% stratum had 2.07 OR (95% CI [1.90,2.23]) relative to age 17, and individuals at the 10-50% stratum had 1.76 OR (95% CI [1.63,1.91]) relative to age 17. Individuals at the bottom stratum, 0-10%, had 1.43 the OR (95% CI [1.23, 1.67]). In contrast, in Whites, the ORs at age 50 relative to age 17 are 1.43, 1.27, 1.22, 1.15 for the 90-100%, 50-90%, 10-50%, and 1-10% strata, respectively.

### PRS association with disease outcomes in MGB Biobank

Figure S9 in the Supplementary Materials describes the association of the multi-ethnic hypertension PRS with hypertension, ischemic stroke, CAD, type 2 diabetes, CKD, and obesity, in the MGB Biobank. In multi-ethnic analysis, the PRS was associated with hypertension (OR=1.45, p-value< 9×10^-100^), as well as with all other outcomes (OR =1.1-1.4 for all outcomes, p-value<0.05). Association of the HTN-PRS with obesity is likely because the pan-ancestry UKBB GWAS of hypertension, which we used, was not adjusted for BMI, and/or due to residual effects of BMI that were not fully accounted for by the BMI adjustment in the MVP GWAS.

## Discussion

We developed a HTN-PRS based on multi-ethnic GWAS for SBP, DBP and assessed its association in a multi-ethnic TOPMed dataset. A BioMe stage 1 dataset was used to select optimal tuning parameters for PRS based on each GWAS using three approaches: the novel Selected CV-PRS approach, Selected PVAL-PRS, and genome-wide significant PRS. We further proposed to combine BP phenotypes PRS based on GWAS of different phenotypes using the PRSsum approach: an unweighted sum of the separate phenotypes’ PRS. This final HTN-PRS, PRSsum based on Selected CV-PRS, was associated with hypertension prevalence in the independent stage 2 dataset, as well as with longitudinal categories of BP trajectories across race/ethnic backgrounds. In analysis stratified by age decade, the association of the PRS with both prevalent and incident hypertension is consistent across ages 21 to 80. In the stage 3 CARDIA study of young adults with 15-years follow up, individuals in strata defined by the top decile of the PRS developed hypertension earlier, especially Black individuals, who tend to have higher PRS values compared to White individuals. Thus, the HTN-PRS can be potentially useful for assessing risk for developing hypertension throughout adulthood. Finally, the HTN-PRS was significantly associated with cardiovascular outcomes in the MGB Biobank (stage 4 dataset).

Recently, a study in Finnish Europeans from FinnGen (26), studied the use of BP PRS to predict longitudinal and lifetime risk of hypertension. The PRS were highly associated hypertension and with cardiovascular disease (CVD) risk, underscoring the potential of PRS to predict hypertension and stratify individuals for intervention to potentially reduce CVD risk. Here, we addressed a similar problem while focusing on a multi-ethnic population and on 4-6 years from between two exams. The distribution of the various constructed PRS, including the final one, PRSsum based on Selected CV-PRS, differed across race/ethnic backgrounds. This is expected, because PRS are sums of alleles, which have different distributions (defined by allele frequencies) across genetic ancestries, and therefore, also race/ethnic background, as these generally have different ancestry admixture. While we expect PRS values in the upper decile to be associated with higher risk of hypertension across all race/ethnicities, a natural question is how to define individuals as “at risk”. An “at risk” classification may use a specific cut-off value of the PRS, which may be based on a percentile of the distribution (42). Clearly, this approach cannot be used when distributions differ across race/ethnicities, and moreover, admixed individuals are not accurately represented by any specific distribution. Therefore, more work is needed on approaches that do not require categorization of neither individuals nor of specific PRS values to define risk. Rather, models that take into account multiple risk factors (such as demographic, clinical, and other risk factors, as well as PRS (43)) and allow for flexible association model may be more powerful and equitable, in that they could be applied to more individuals. Notably, we could not use the standard approach of quantifying hypertension risk between individuals in the top HTN-PRS decile to the bottom, in the multi-ethnic analysis: such an approach would separate AAs from others, and therefore will be confounded by other social race/ethnic-related environmental exposures.

Methodologically, while we first constructed various PRS using a standard clump and threshold methodology (22), we used two novel approaches to construct the new HTN-PRS. First, we leveraged stage 1 and stage 2 independent datasets to study how to select the tuning parameters for a PRS, and chose the Selected CV-PRS as a method. This approach attempts to avoid overfitting to a particular dataset by splitting the training data to 5 independent subsets, i.e. with no related individuals between them, and assessing association of the PRS with the outcome in each. The Selected CV-PRS is the one that has consistent, high, effect size across these subsets, represented by smallest CV across them. Other measures can be used rather than effect size, but we chose the latter because of clinical interpretability. In the testing dataset, the effect size was indeed the highest when using the Selected CV-PRS compared with other PRS. A second methodological choice was the construction of PRSsum. PRSsum allowed us to combine information across PRS that were based on GWAS of different phenotypes (SBP, DBP, hypertension). GWAS of different phenotypes cannot be meaningfully meta-analyzed to produce new summary statistics for PRS construction. Another motivation behind it is that high values of PRSsum may capture individuals with either high SBP, DBP, or hypertension PRS values, or combined, meaning that their hypertension may be captured by various underlying genetic components. While PRSsum is an unweighted sum of PRS, a weighted sum (or with adaptive weights) can be constructed as well (44, 45). We here used PRSsum without adaptive weights out of concern for overfitting due to a dominant PRS and dominant subgroup, for example, where the PRS based on hypertension GWAS would potentially be upweighted due to a large set of individuals with European ancestry. Future work should study weighted combination of PRS in diverse populations.

Strengths of our study are the use of large multi-ethnic datasets with harmonized genetic and phenotypic data, a range of ages of participants, and longitudinal datasets, allowing us to explore the association of hypertension PRS across adulthood. In stage 3, we focused on one study, CARDIA, with longer follow up of younger individuals, and compared trajectories of hypertension risk by age across Black and White individuals, demonstrating that Blacks develop hypertension early, in agreement with their higher PRS values. Additional longitudinal datasets from underrepresented populations are needed to study long-term trajectories of disease development and usefulness of PRS across the lifespan. Our study also has additional weaknesses. For example, our primary analysis did not use the largest available GWAS of SBP and DBP to construct PRS, namely a meta-analysis of the European ancestry participants UKBB and of the international consortium for BP (9) as most of our study individuals participated in it, and the overlap could lead to overfitting. More work is needed to assess overfitting effects across samples sizes and overlaps of discovery GWAS, training, and testing datasets. Also, to construct PRS we used the clump & threshold methodology, rather than a more modern approach such as LDpred (46) or lassosum (47). We chose to focus on clump & threshold methodology because we think that these other methods still need to be separately studied for diverse populations.

In summary, we applied novel methodology for developing PRS and constructed a PRS predictive of incident hypertension across adulthood in a multi-ethnic population. The PRS was also significantly associated with clinical outcomes. Future work will incorporate rare variants and pleiotropic variants (48) in the construction of PRS, and will investigate models for clinical uses of hypertension PRS in diverse populations.

## Data Availability

TOPMed freeze 8 WGS data are available by application to dbGaP according to the study specific accessions:  ARIC: phs001416, BioMe: phs001644, CFS: phs000954, CHS: phs001368, FHS: phs000974, GENOA: phs001345, HCHS/SOL: phs001395, JHS: phs000964, MESA: phs001211, WHI: phs001237. Study phenotypes are available from dbGaP from study accession: ARIC: phs000090, BioMe: phs001644, CFS: phs000284, CHS: phs000287, FHS: phs000007, GENOA: phs000379, HCHS/SOL: phs000810, JHS: phs000286, MESA: phs000209, WHI: phs000200. Data needed to construct the HTN-PRS will be provided upon paper acceptance as a data supplement, and includes variants, alleles, and weights for each of the Selected CV-PRS based on GWAS of SBP, DBP, and hypertension, mean and SD computed based on the TOPMed-BP dataset, and code to generate the multi-ethnic PRS from plink files using PRSice 2. They are also publicly available on the repository https://github.com/nkurniansyah/Hypertension_PRS. 

## Acknowledgements

We gratefully acknowledge the studies and participants who provided biological samples and data for TOPMed and CCDG. TOPMed and CCDG acknowledgements, as well as descriptions, acknowledgements, and ethics statements of contributing studies are provided in the Supplementary Materials. The views expressed in this manuscript are those of the authors and do not necessarily represent the views of the National Heart, Lung, and Blood Institute; the National Institutes of Health; or the U.S. Department of Health and Human Services.

## Declaration of Interests

B Psaty serves on the Steering Committee of the Yale Open Data Access Project funded by Johnson & Johnson. All other co-authors declare no conflict of interest.

## Supplementary Tables

**Table S1:**
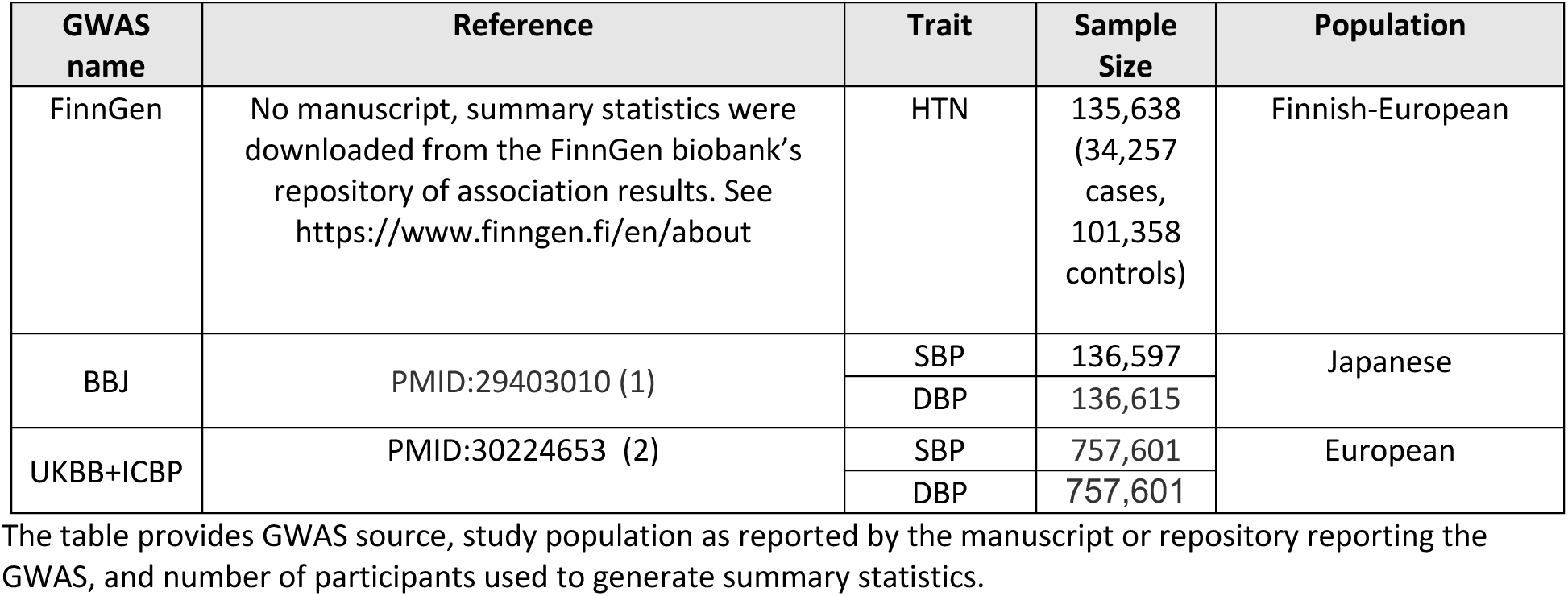
External GWAS used for hypertension PRS construction in secondary analysis

**Table S2:** Characteristics of the TOPMed stage 1 (BioMe) training dataset

| Characteristics | African-American | Asian-American | European-American | Hispanic-American | Others |
| --- | --- | --- | --- | --- | --- |
| N | 2803 | 151 | 2506 | 4187 | 667 |
| Sex (%) F | 1864 (66.5) | 84 (55.6) | 1198 (47.8) | 2616 (62.5) | 281 (42.1) |
| Age (mean (SD)) | 55.99 (12.50) | 53.14 (13.52) | 60.42 (13.96) | 57.72 (13.18) | 57.51 (13.10) |
| Current smoke (%) Yes | 658 (23.6) | 10 ( 6.6) | 240 ( 9.6) | 741 (17.8) | 94 (14.2) |
| Antihypertensive medication (%) Yes | 1589 (56.7) | 48 (31.8) | 807 (32.2) | 2200 (52.5) | 255 (38.2) |
| BMI (mean (SD)) | 30.46 (8.20) | 24.30 (3.85) | 26.58 (7.36) | 29.24 (6.50) | 27.61 (10.35) |
| SBP (mean (SD)) | 145.03 (28.57) | 129.93 (27.36) | 130.34 (23.75) | 139.36 (28.04) | 133.38 (24.78) |
| DBP (mean (SD)) | 82.42 (15.76) | 73.71 (14.97) | 73.68 (13.14) | 78.04 (14.53) | 75.79 (13.39) |
| Hypertension (%) Yes | 2222 (79.3) | 85 (56.3) | 1477 (58.9) | 3084 (73.7) | 420 (63.0) |

**Table S3:** Characteristics of TOPMed stage 2 dataset participants.

| Characteristics | African-American | Asian-American | European-American | Hispanic-American |
| --- | --- | --- | --- | --- |
| N | 8822 | 794 | 22701 | 6718 |
| Sex (%) F | 6004 (68.1) | 493 (62.1) | 16395 (72.2) | 4168 (62.0) |
| Cohort (%) |  |  |  |  |
| • ARIC | 1324 (15.0) | 0 ( 0.0) | 6000 (26.4) | 0 ( 0.0) |
| • CHS | 692 ( 7.8) | 0 ( 0.0) | 2765 (12.2) | 24 ( 0.4) |
| • FHS | 0 ( 0.0) | 0 ( 0.0) | 3102 (13.7) | 11 ( 0.2) |
| • GENOA | 1069 (12.1) | 0 ( 0.0) | 0 ( 0.0) | 0 ( 0.0) |
| • HCHS_SOL | 0 ( 0.0) | 0 ( 0.0) | 0 ( 0.0) | 5357 (79.7) |
| • JHS | 3211 (36.4) | 0 ( 0.0) | 0 ( 0.0) | 0 ( 0.0) |
| • MESA | 1096 (12.4) | 599 (75.4) | 1856 ( 8.2) | 1019 (15.2) |
| • WHI | 1430 (16.2) | 195 (24.6) | 8978 (39.5) | 307 ( 4.6) |
| Year between visit (mean (SD)) | 4.66 (1.03) | 4.45 (0.83) | 4.81 (1.62) | 5.71 (1.00) |
| Age at baseline (mean (SD)) | 58.72 (11.29) | 62.44 (9.78) | 59.97 (12.73) | 50.76 (13.46) |
| Current smoke at baseline (%) Yes | 1630 (18.5) | 37 ( 4.7) | 4463 (19.7) | 1310 (19.5) |
| Antihypertensive medication at baseline (%) Yes | 4637 (52.8) | 239 (30.1) | 6908 (30.5) | 1532 (23.0) |
| BMI at baseline (mean (SD)) | 30.92 (6.69) | 24.29 (3.62) | 27.34 (5.34) | 30.06 (6.10) |
| SBP at baseline (mean (SD)) | 139.20 (22.58) | 131.18 (24.70) | 130.80 (22.25) | 127.64 (21.29) |
| DBP at baseline (mean (SD)) | 82.24 (11.61) | 76.90 (12.09) | 76.75 (11.35) | 76.51 (11.84) |
| Hypertension at baseline (%) Yes | 6711 (76.5) | 452 (57.5) | 13546 (60.1) | 3422 (51.7) |
| Antihypertensive medication at follow-up(%) Yes | 4767 (66.1) | 305 (41.8) | 8146 (39.4) | 2575 (39.5) |
| BMI at follow-up (mean (SD)) | 31.19 (6.74) | 24.29 (3.68) | 27.82 (5.55) | 30.31 (6.20) |
| SBP at follow-up (mean (SD)) | 141.33 (22.28) | 131.41 (23.29) | 132.07 (21.73) | 130.80 (22.64) |
| DBP at follow up (mean (SD)) | 81.71 (11.50) | 75.85 (11.25) | 76.25 (11.08) | 77.12 (11.97) |
| Hypertension at follow-up (%) Yes | 5972 (83.3) | 438 (60.8) | 13034 (63.8) | 3860 (59.6) |

**Table S4.** Tuning parameters and SNP counts in primary and secondary PRS trained using stage 1 dataset

| Genome-wide Significant |  |  |  |  |
| --- | --- | --- | --- | --- |
| Diastolic BP |  |  |  |  |
| Study | Threshold | R2 | Distance | # SNP |
| BBJ | 5.00E-08 | 0.1 | 1000kb | 46 |
| MVP |  |  |  | 86 |
| Pan UKBB |  |  |  | 1319 |
| UKBB+ICBP |  |  |  | 1730 |
| BBJ+MVP+Pan UKBB |  |  |  | 101 |
| Systolic BP |  |  |  |  |
| BBJ | 5.00E-08 | 0.1 | 1000kb | 67 |
| MVP |  |  |  | 144 |
| Pan UKBB |  |  |  | 1235 |
| UKBB+ICBP |  |  |  | 1602 |
| BBJ+MVP+Pan UKBB |  |  |  | 166 |
| Hypertension |  |  |  |  |
| FINNGEN | 5.00E-08 | 0.1 | 1000kb | 15 |
| Pan UKBB |  |  |  | 582 |
| FINNGEN+Pan UKBB |  |  |  | 382 |
| Selected CV-PRS |  |  |  |  |
| Diastolic BP |  |  |  |  |
| BBJ | 5.00E-01 | 0.3 | 250kb | 245160 |
| MVP | 1.00E-01 | 0.3 | 250kb | 149559 |
| Pan UKBB | 4.00E-01 | 0.1 | 1000kb | 291655 |
| UKBB+ICBP | 2.00E-01 | 0.1 | 1000kb | 131193 |
| BBJ+MVP+Pan UKBB | 2.00E-01 | 0.1 | 250kb | 253855 |
| Systolic BP |  |  |  |  |
| BBJ | 1.00E-02 | 0.2 | 1000kb | 10280 |
| MVP | 1.00E-02 | 0.2 | 1000kb | 24807 |
| Pan UKBB | 1.00E-05 | 0.1 | 1000kb | 3291 |
| UKBB+ICBP | 1.00E-01 | 0.1 | 1000kb | 87939 |
| BBJ+MVP+Pan UKBB | 5.00E-01 | 0.3 | 2500kb | 444645 |
| Hypertension |  |  |  |  |
| FINNGEN | 1.00E-02 | 0.1 | 500kb | 14215 |
| Pan UKBB | 3.00E-01 | 0.1 | 250kb | 234228 |
| FINNGEN+Pan UKBB | 1.00E-04 | 0.1 | 1000kb | 2486 |

| **Selected PVAL-PRS** |  |  |  |  |
| --- | --- | --- | --- | --- |
| ***Diastolic BP*** |  |  |  |  |
| BBJ | 5.00E-01 | 0.3 | 500kb | 243785 |
| MVP | 1.00E-01 | 0.3 | 1000kb | 147123 |
| Pan UKBB | 1.00E-01 | 0.2 | 250kb | 186307 |
| UKBB+ICBP | 1.00E-02 | 0.1 | 1000kb | 28354 |
| BBJ+MVP+Pan UKBB | 1.00E-01 | 0.1 | 1000kb | 15064 |
| ***Systolic BP*** |  |  |  |  |
| BBJ | 2.00E-01 | 0.3 | 500kb | 122815 |
| MVP | 1.00E-03 | 0.1 | 1000kb | 4643 |
| Pan UKBB | 1.00E-02 | 0.1 | 1000kb | 40148 |
| UKBB+ICBP | 1.00E-02 | 0.1 | 500kb | 28613 |
| BBJ+MVP+Pan UKBB | 1.00E-04 | 0.3 | 250kb | 1710 |
| ***Hypertension*** |  |  |  |  |
| FINNGEN | 1.00E-02 | 0.3 | 1000kb | 18937 |
| Pan UKBB | 2.00E-01 | 0.3 | 250kb | 302815 |
| FINNGEN+Pan UKBB | 1.00E-04 | 0.1 | 500kb | 2535 |

**Table S5.** Characteristics of the stage 3 CARDIA dataset

|  | Baseline |  | Follow up visit I<br>(2 years) |  | Follow up visit II<br>(5 years) |  | Follow up visit III<br>(7 years) |  | Follow up visit IV<br>(10 years) |  | Follow up visit V<br>(15 years) |  |
| --- | --- | --- | --- | --- | --- | --- | --- | --- | --- | --- | --- | --- |
| Characteristics | Black | White | Black | White | Black | White | Black | White | Black | White | Black | White |
| N | 1388 | 1699 | 1268 | 1641 | 1258 | 1634 | 1221 | 1591 | 1246 | 1571 | 1239 | 1602 |
| Sex (%) F | 836<br>(60.2) | 918<br>(54.0) | 765<br>(60.3) | 885<br>(53.9) | 765<br>(60.2) | 878<br>(54.0) | 748<br>(61.3) | 848<br>(53.3) | 757<br>(60.8) | 842<br>(53.6) | 751<br>(60.6) | 865<br>(54.0) |
| Age (mean (SD)) | 24.44<br>(3.81) | 25.67<br>(3.31) | 26.43<br>(3.80) | 27.71<br>(3.29) | 29.51<br>(3.79) | 30.71<br>(3.30) | 31.51<br>(3.80) | 32.68<br>(3.29) | 34.47<br>(3.80) | 35.68<br>(3.31) | 39.49<br>(3.83) | 40.70<br>(3.30) |
| BMI (mean (SD)) | 25.46<br>(5.80) | 23.61<br>(3.95) | 26.40<br>(6.21) | 24.22<br>(4.22) | 27.60<br>(6.60) | 24.86<br>(4.59) | 28.34<br>(6.77) | 25.47<br>(5.03) | 29.14<br>(7.01) | 25.94<br>(5.25) | 30.67<br>(7.55) | 27.18<br>(5.85) |
| SBP (mean (SD)) | 111.43<br>(11.31) | 109.11<br>(10.93) | 109.80<br>(11.80) | 106.45<br>(10.61) | 110.04<br>(12.44) | 105.58<br>(10.95) | 111.48<br>(13.33) | 106.18<br>(11.53) | 113.23<br>(14.26) | 107.18<br>(11.40) | 118.71<br>(17.98) | 110.37<br>(13.74) |
| DBP (mean (SD)) | 68.88<br>(9.98) | 68.40<br>(9.22) | 68.94<br>(10.41) | 66.97<br>(9.15) | 70.98<br>(11.06) | 67.74<br>(9.55) | 71.05<br>(11.11) | 67.89<br>(9.65) | 74.94<br>(11.15) | 70.46<br>(9.38) | 78.06<br>(13.40) | 72.67<br>(10.58) |
| Antihypertensive<br>medication (%) Yes | 17<br>(1.2) | 11<br>(0.6) | 46<br>(3.6) | 24<br>(1.5) | 35<br>(2.8) | 11<br>(0.7) | 36<br>(3.0) | 17<br>(1.1) | 70<br>(5.6) | 22<br>(1.4) | 163<br>(13.2) | 65<br>(4.1) |
| Current Smoke (%)<br>Yes | 405<br>(29.7) | 403<br>(24.1) | 494<br>(39.0) | 514<br>(31.3) | 423<br>(33.6) | 436<br>(26.7) | 862<br>(70.6) | 1277<br>(80.3) | 884<br>(70.9) | 1268<br>(80.7) | 895<br>(72.2) | 1328<br>(82.9) |
| Hypertension (%)<br>Yes | 214<br>(15.4) | 211<br>(12.4) | 207<br>(16.3) | 140<br>(8.5) | 254<br>(20.2) | 175<br>(10.7) | 272<br>(22.3) | 178<br>(11.2) | 369<br>(29.7) | 256<br>(16.3) | 518<br>(41.9) | 389<br>(24.3) |

## Supplementary figures

**Figure S1:**
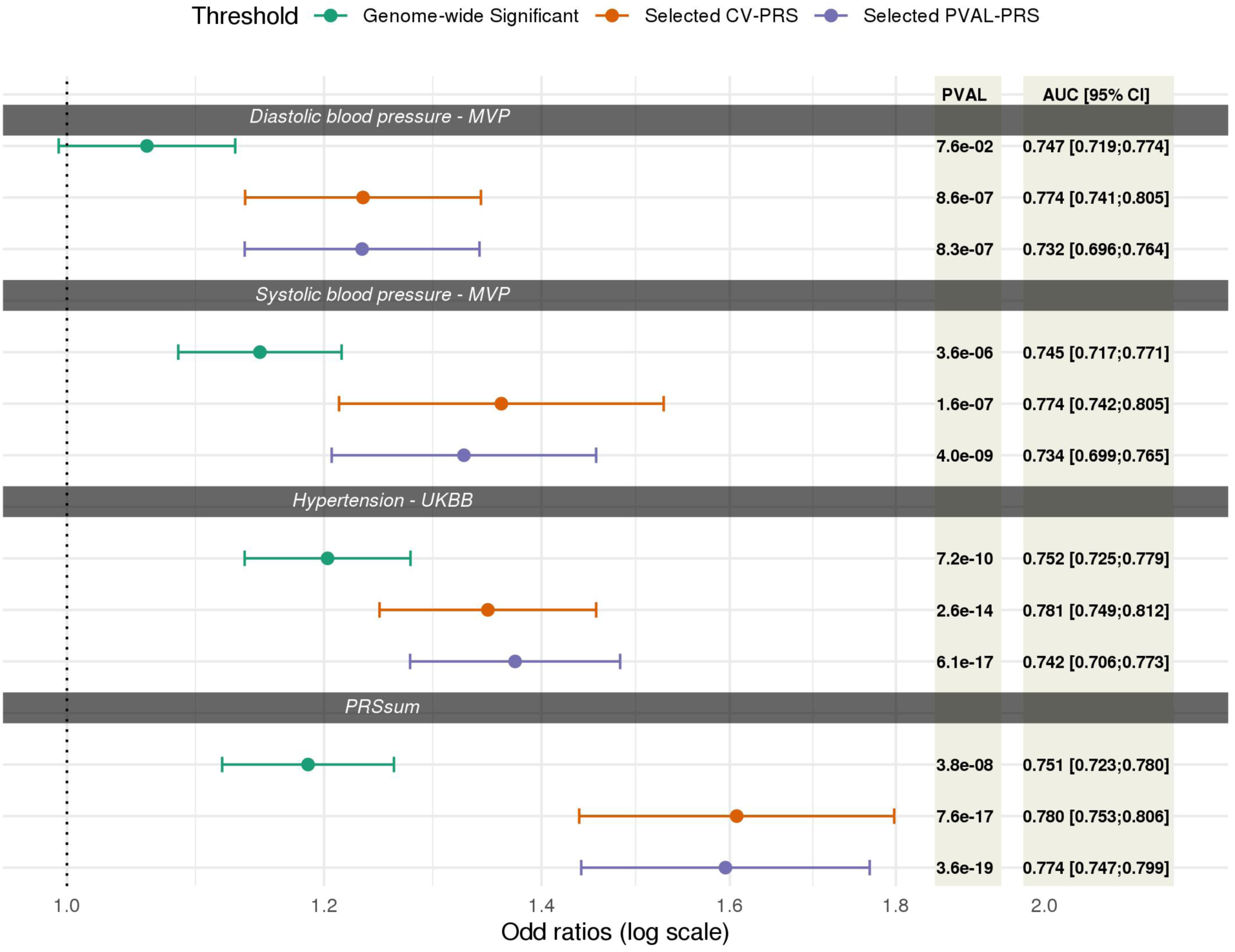
Association of primary PRS with hypertension in the stage 1 dataset across compared tuning parameter selection criteria. For each GWAS, the PRS were selected based on: (1) Genome wide significant SNPs, using SNPs with p-value<5×10^-8^, and fixed clumping parameters: R^2^=0.1 and distance of 1000kb, (2) Optimizing the coefficient of variation across 5 independent subsets of the stage 1 dataset (see Figure 1), and (3) The PRS with the lowest association P-value. We provide OR, 95% confident interval, association P-value and AUC. The PRS associations were estimated in models adjusted for sex, age, age^2^, study site, race/ethnic background, smoking status, BMI, and 11 ancestral principal components.

**Figure S2.**
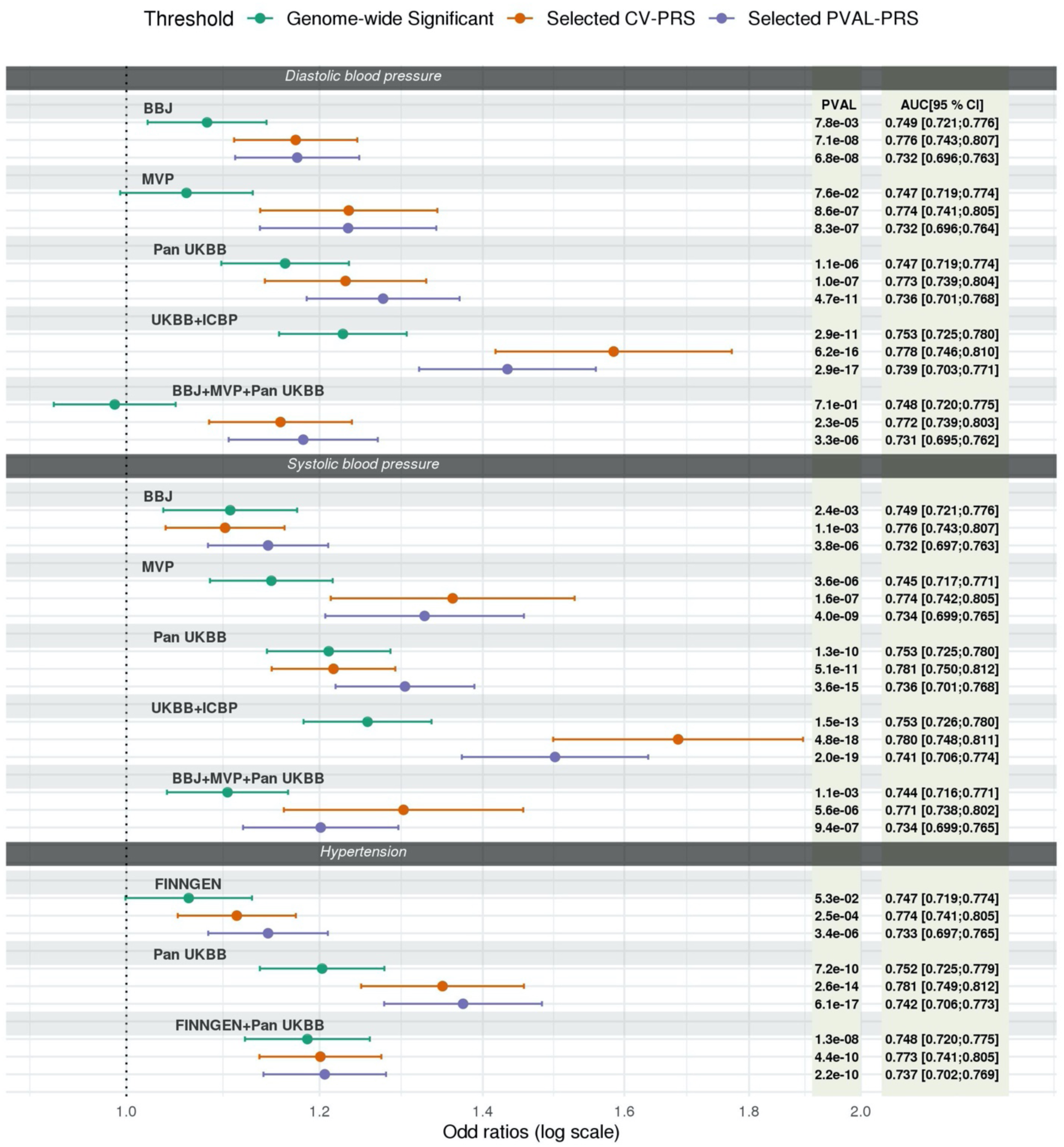
Association of primary and secondary PRS with hypertension in the BioMe stage 1 dataset across tuning parameter selection criteria. For each of the primary and secondary GWAS used, the PRS were selected based on: (1) Genome wide significant SNPs, using SNPs with p-value<5×10^-8^, and fixed clumping parameters: R^2^=0.1 and distance of 1000kb,(2) Optimizing the coefficient of variation across 5 independent subsets of the stage 1 dataset (see Figure 1), and (3) The PRS with the lowest association P-value. We provide OR, 95% confident interval, association p-value and AUC. The PRS associations were estimated in models adjusted for sex, age, age^2^, study site, race/ethnic background, smoking status, BMI, and 11 ancestral principal components.

**Figure S3.**
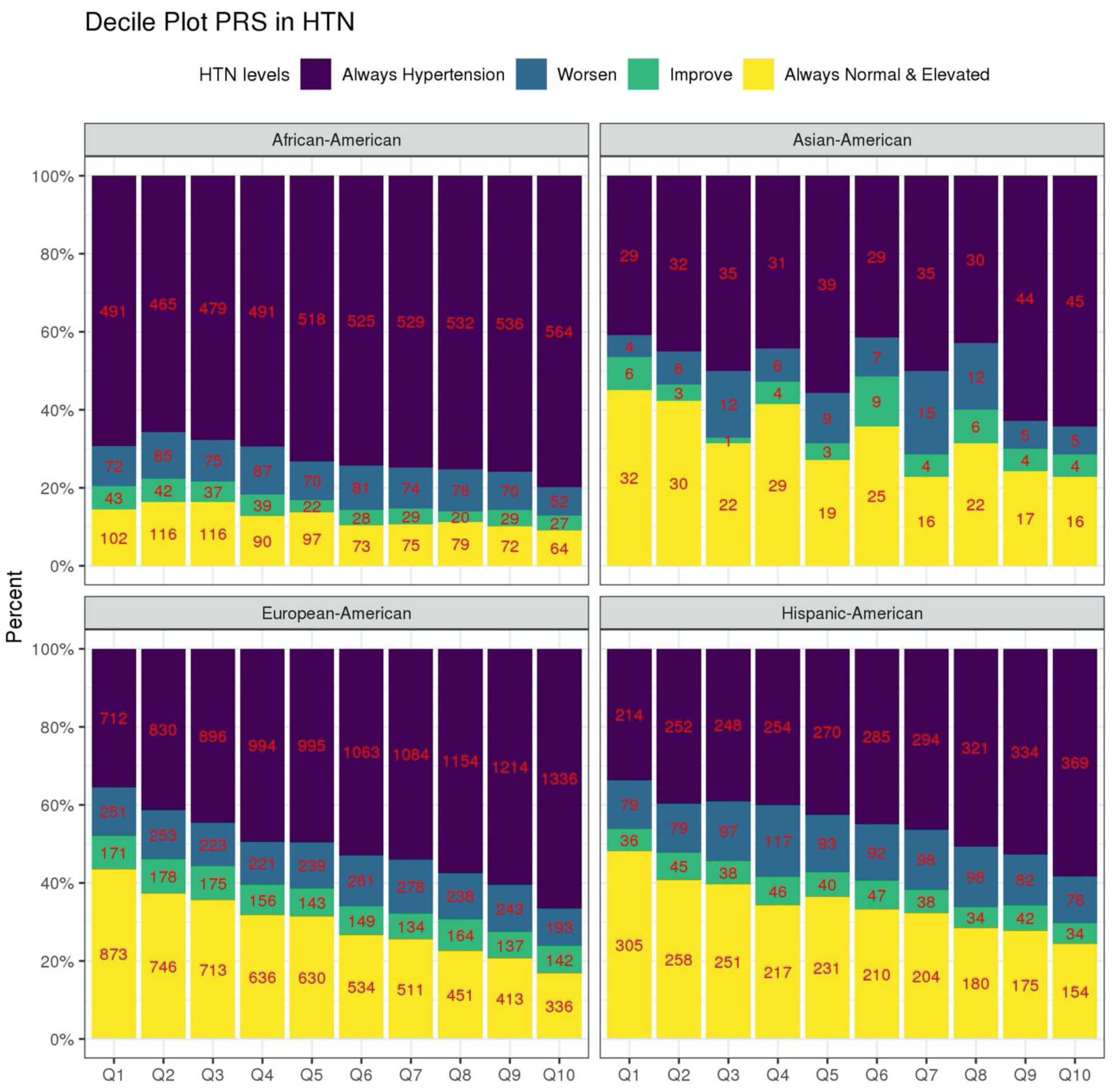
Distribution of longitudinal categories of BP stratified by race/ethnic background. The figure visualizes the distribution of hypertension severity measures stratified by race/ethnic background. Severity measures from most to least severe: always hypertension (treated and hypertension in both visit), worsen (change from normal to elevated BP or hypertension, or change from elevated to hypertension) improve (change from untreated hypertension at to elevated/normal at follow up, or change from elevated to normal), no hypertension in both exams in deciles of the HTN-PRS (PRSsum based on Selected CV-PRS). The numbers provide the sample sizes represented by each bar.

**Figure S4.**
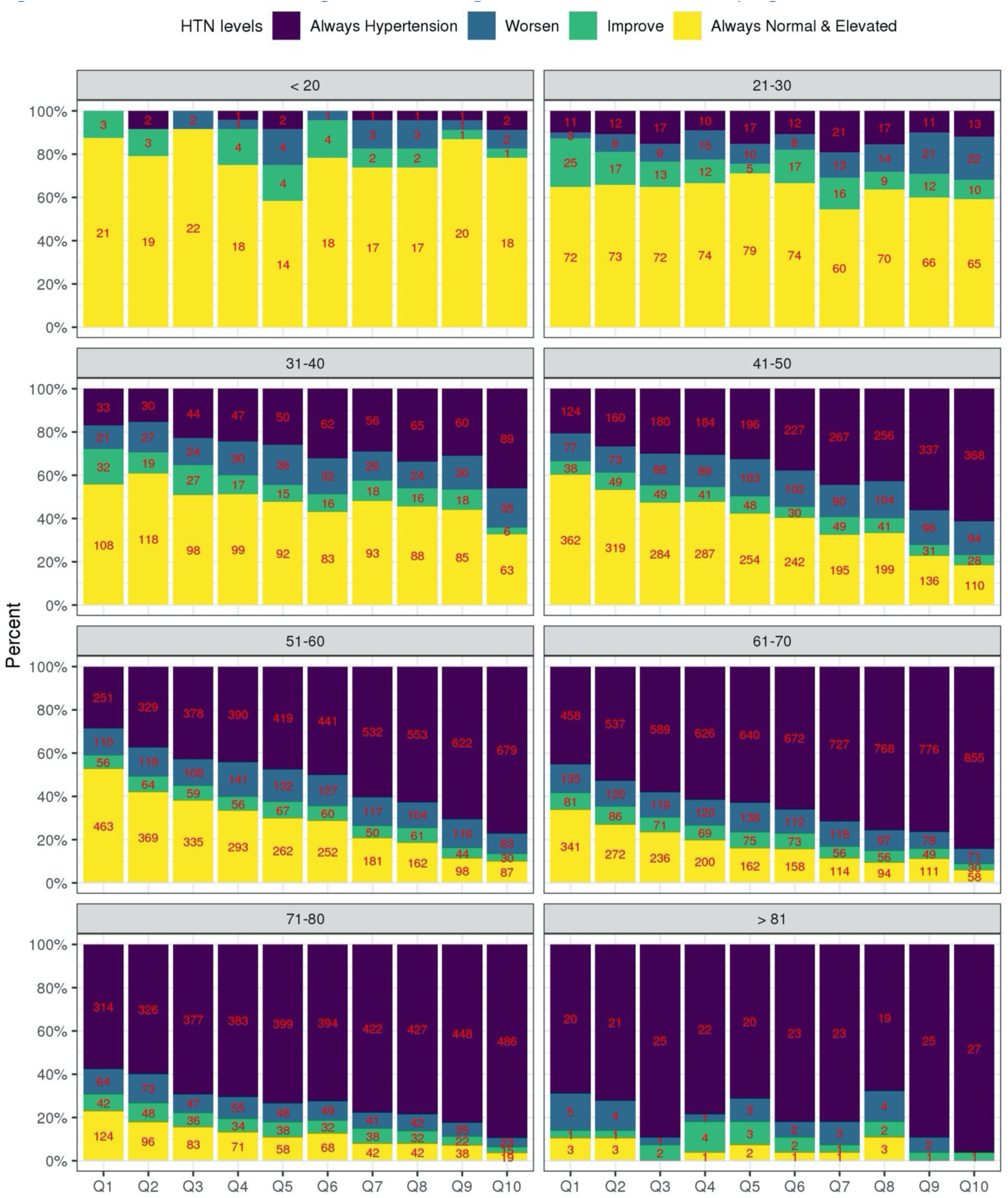
Distribution of longitudinal categories of BP stratified by age. The figure visualizes the distribution of hypertension severity measures stratified by age (age (≤20, 21-30, 31-40, … 71-80, >80). Severity measures from most to least severe: always hypertension (treated and hypertension in both visit), worsen (change from normal to elevated BP or hypertension, or change from elevated to hypertension) improve (change from untreated hypertension at to elevated/normal at follow up, or change from elevated to normal), no hypertension in both exams in deciles of the HTN-PRS (PRSsum based on Selected CV-PRS). The numbers provide the sample sizes represented by each bar.

**Figure S5:**
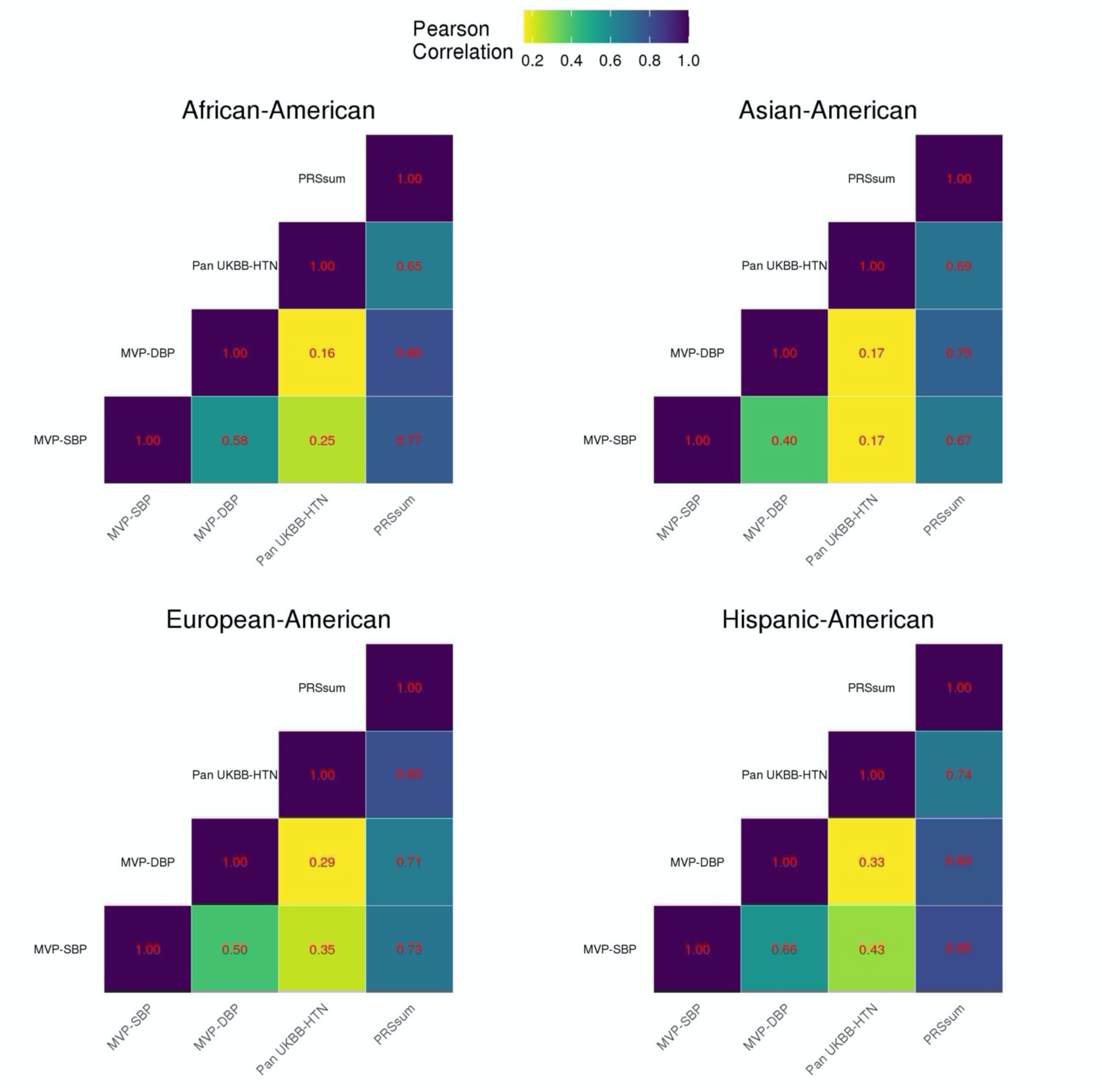
Correlation heatmap between Selected CV PRS by phenotype and PRSsum stratified by race/ethnic background. Correlations were computed using the stage 2 dataset.

**Figure S6.**
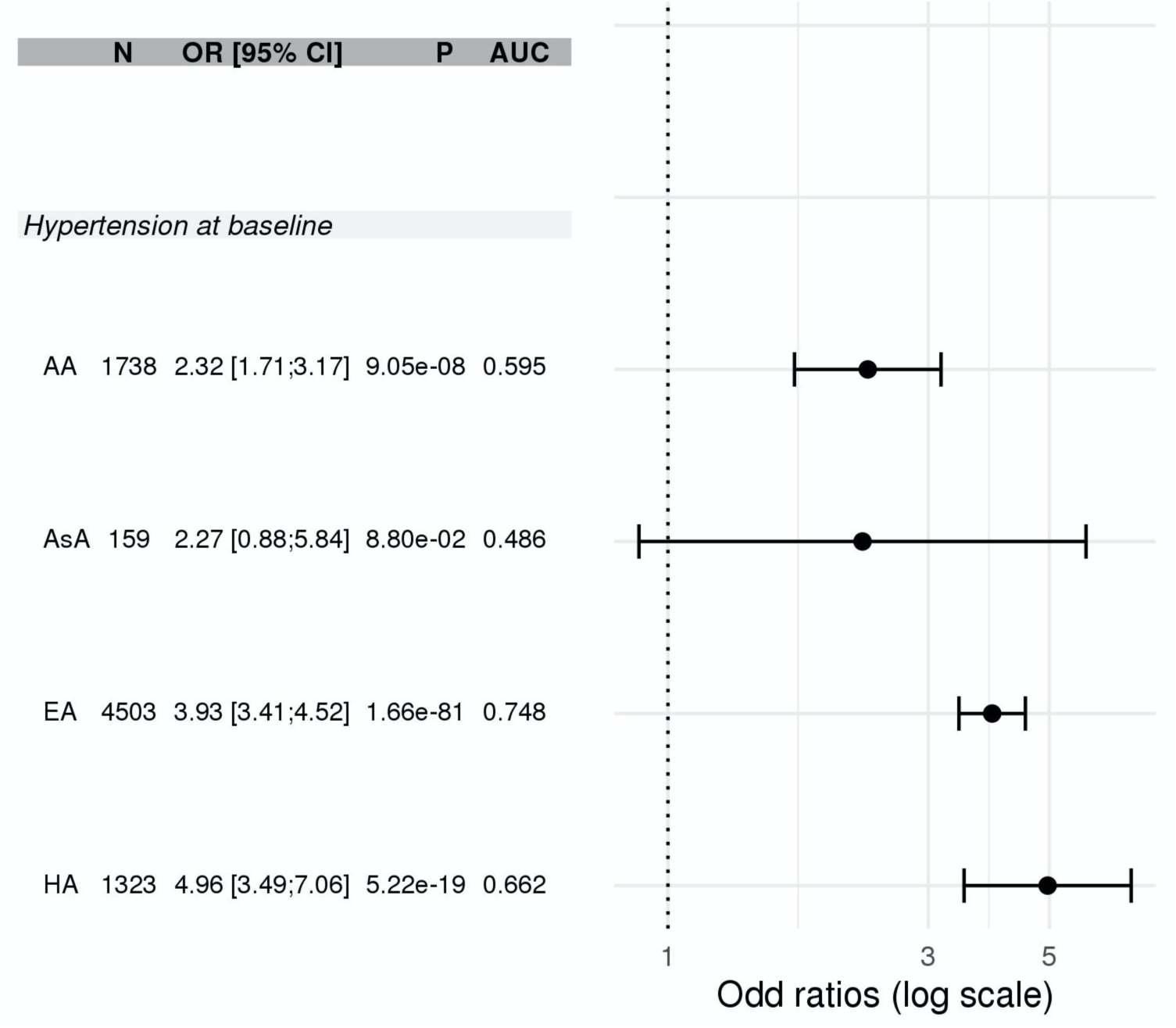
Risk of hypertension in individuals in high deciles of the HTN-PRS compared to those in low deciles of the HTN-PRS. The forest plot provides OR for hypertension at baseline in individuals in the highest versus the lowest decile of the HTN-PRS. Associations were computed in the stage 2 dataset, and are provided in each race/ethnic group separately, because the PRS distribution differ between race/ethnicities.

**Figure S7.**
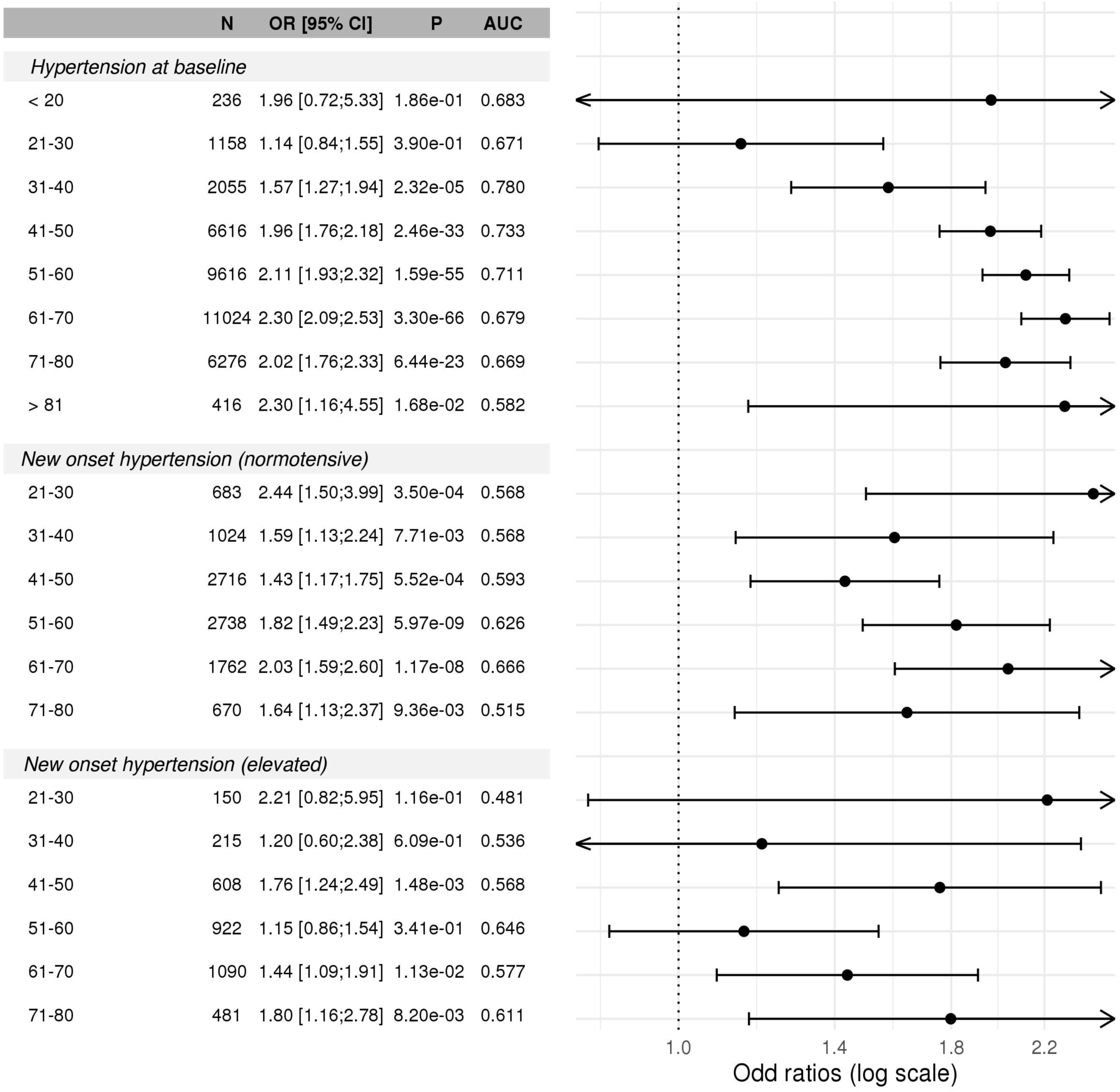
Age-stratified associations of HTN-PRS with prevalent and incident hypertension. The forest plot provides the association of the HTN-PRS with prevalent and incident hypertension in the stage 2 dataset, and within age groups. Incident hypertension analysis was performed within individuals who had normal BP at baseline (normotensive), and within individuals who had elevated BP at baseline (elevated). For each analysis we provide sample size, estimated odds ratio (OR) and 95% confidence interval, p-value, and area under the receiver operating curve (AUC).

**Figure S8:**
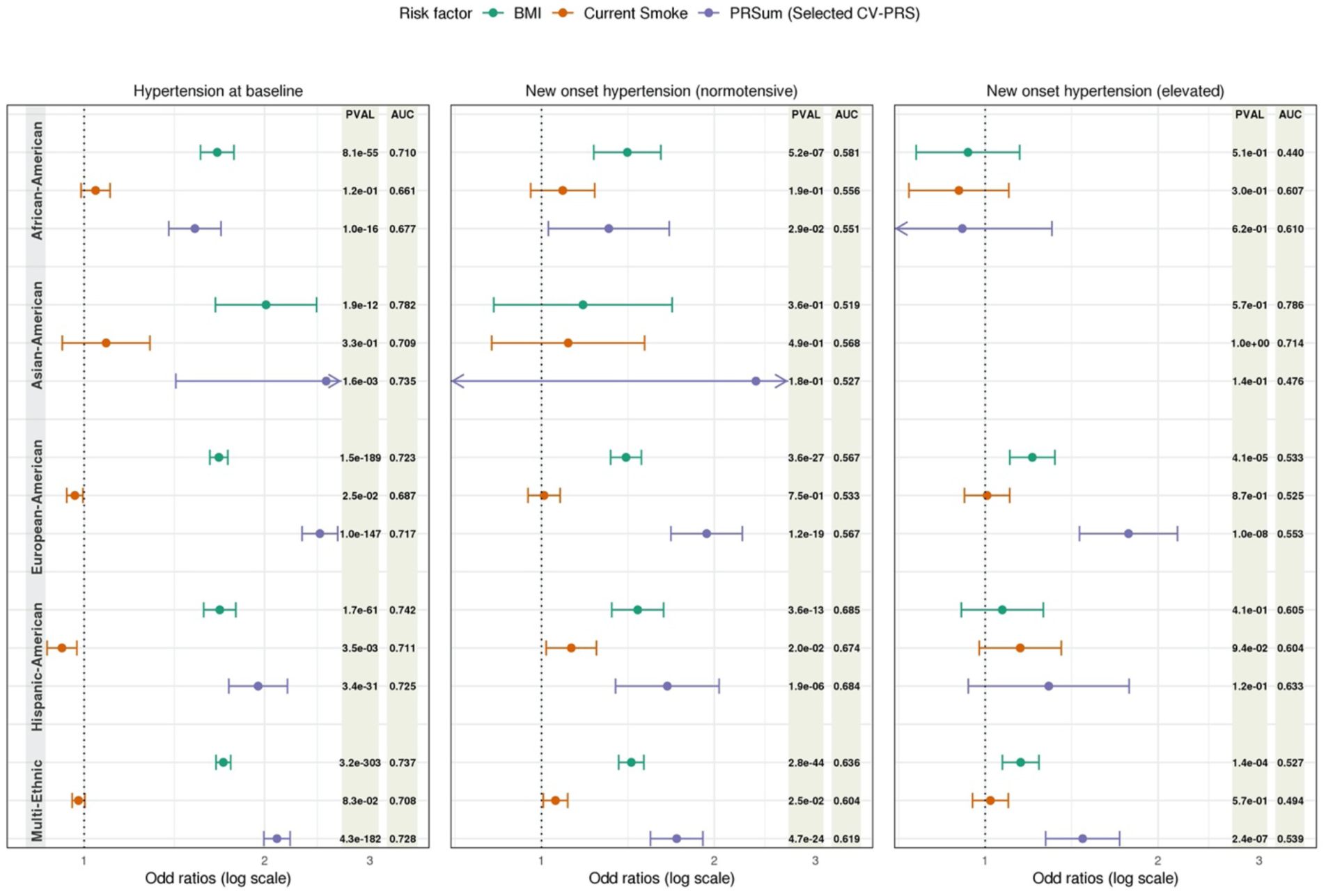
Benchmarking the risk of HTN-PRS against other risk factors. The figure compares hypertension risk factors: BMI, current smoking, and HTN-PRS. For each association analysis, the figure provides the within-group standardized effect size estimate (effect size per 1SD increase in the risk factor within the race/ethnic group used) and 95% confidence interval. We also provide the AUC from a model that include covariates (sex, age, age^2^, study site, race/ethnicity in the multi-ethnic model, and 11 PCs) and only the single risk factor of interest (and not the other two risk factors).

**Figure S9:**
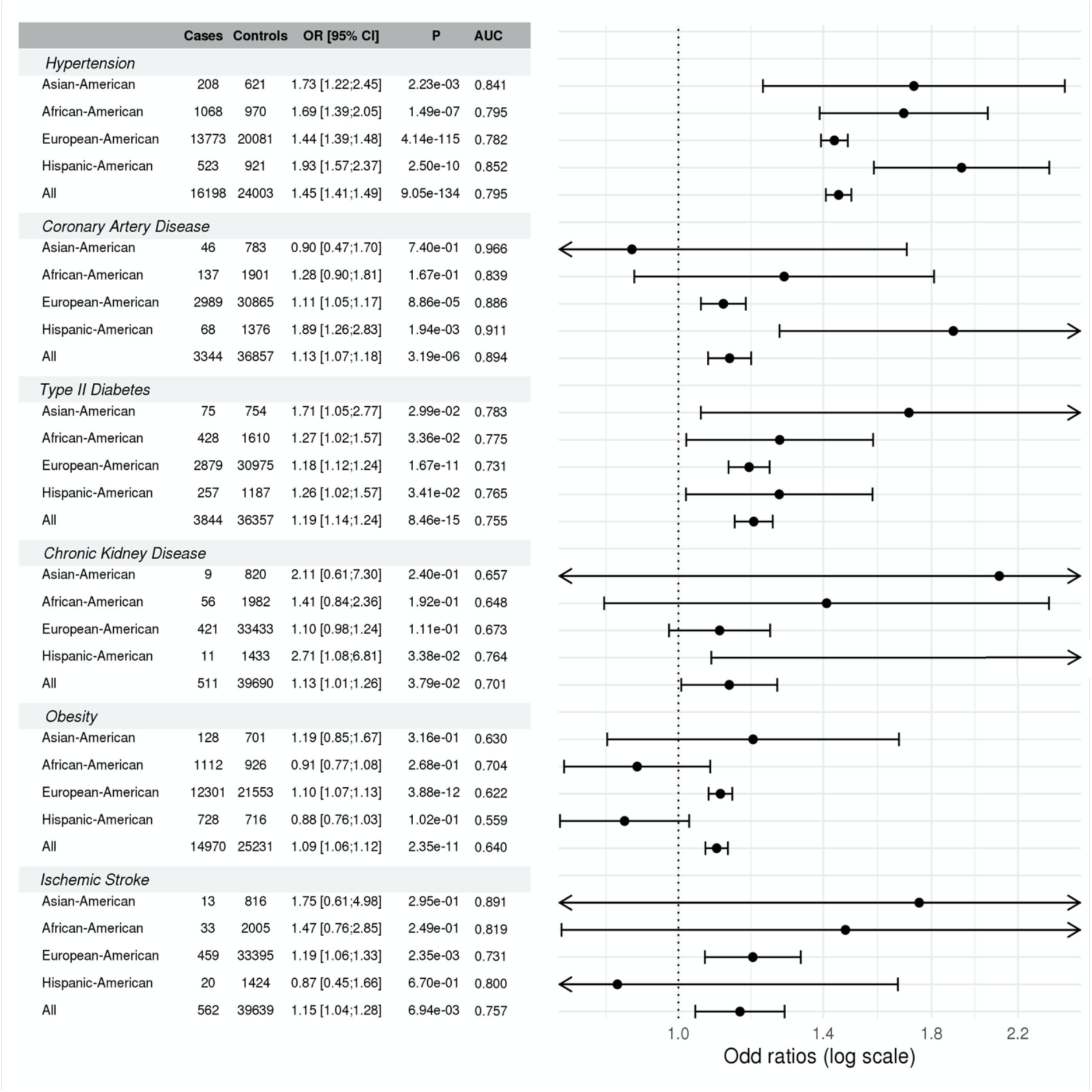
Race/ethnicity stratified associations of the HTN-PRS with outcomes in the MGB Biobank. The figure provides the association of HTN-PRS with outcomes in the MGB Biobank, stratified by race/ethnic background groups.

## Study descriptions

### BioMe

The BioMe Biobank is an ongoing, prospective, hospital- and outpatient- based population research program operated by The Charles Bronfman Institute for Personalized Medicine (IPM) at Mount Sinai. BioMe has enrolled over 50,000 participants between September 2007 and July 2019. BioMe is an Electronic Medical Record (EMR)-linked biobank that integrates research data and clinical care information for consented patients at The Mount Sinai Medical Center, which serves diverse local communities of upper Manhattan with broad health disparities. IPM BioMe populations include 25% of African American ancestry (AA), 36% of Hispanic Latino ancestry (HL), 30% of white European ancestry (EA), and 9% of other ancestry. The BioMe disease burden is reflective of health disparities in the local communities. BioMe operations are fully integrated in clinical care processes, including direct recruitment from clinical sites waiting areas and phlebotomy stations by dedicated BioMe recruiters independent of clinical care providers, prior to or following a clinician standard of care visit. Recruitment currently occurs at a broad spectrum of over 30 clinical care sites.

#### Blood pressure measurements methods

BioMe operations are fully integrated in clinical care processes, including direct recruitment from clinical sites waiting areas and phlebotomy stations by dedicated BioMe recruiters independent of clinical care providers, prior to or following a clinician standard of care visit. Recruitment currently occurs at a broad spectrum of over 30 clinical care sites. Information on anthropometrics, demographics, blood pressure values and use of antihypertensive medication was derived from participants Electronic Medical Record (EMR).

#### Ethics statement

The BioMe cohort was approved by the Institutional Review Board at the Icahn School of Medicine at Mount Sinai. All BioMe participants provided written, informed consent for genomic data sharing.

#### BioMe acknowledgements

The Mount Sinai BioMe Biobank has been supported by The Andrea and Charles Bronfman Philanthropies and in part by Federal funds from the NHLBI and NHGRI (U01HG00638001; U01HG007417; X01HL134588). We thank all participants in the Mount Sinai Biobank. We also thank all our recruiters who have assisted and continue to assist in data collection and management and are grateful for the computational resources and staff expertise provided by Scientific Computing at the Icahn School of Medicine at Mount Sinai.

### ARIC

The Atherosclerosis Risk in Communities (ARIC) study (dbGaP accession phs000090) is a population-based prospective cohort study of cardiovascular disease sponsored by the NHLBI. ARIC included 15,792 individuals, predominantly European American and African American, aged 45-64 years at baseline (1987-89), chosen by probability sampling from four US communities. Cohort members completed three additional triennial follow-up examinations, a fifth exam in 2011-2013, a sixth exam in 2016-2017, and a seventh exam in 2018-2019. The ARIC study has been described in detail previously (3). For this analysis, we used BP measurements from the first and third ARIC exams.

#### Blood pressure measurements methods

BP was measured using a standardized Hawksley random-zero mercury column sphygmomanometer with participants in a sitting position after a resting period of 5 minutes. The size of the cuff was chosen according to the arm circumference. Three sequential recordings for SBP and DBP were obtained; the mean of the last two measurements was used in this analysis, discarding the first reading. Blood pressure lowering medication use was recorded from the medication history

#### Ethics statement

The ARIC study has been approved by Institutional Review Boards (IRB) at all participating institutions: University of North Carolina at Chapel Hill IRB, Johns Hopkins University IRB, University of Minnesota IRB, and University of Mississippi Medical Center IRB. Study participants provided written informed consent at all study visits.

#### ARIC acknowledgements

The Atherosclerosis Risk in Communities study has been funded in whole or in part with Federal funds from the National Heart, Lung, and Blood Institute, National Institutes of Health, Department of Health and Human Services (contract numbers HHSN268201700001I, HHSN268201700002I, HHSN268201700003I, HHSN268201700004I and HHSN268201700005I). The authors thank the staff and participants of the ARIC study for their important contributions.

### CHS

The Cardiovascular Health Study (CHS) is a population-based cohort study initiated by the National Heart, Lung and Blood Institute (NHLBI) in 1987 to determine the risk factors for development and progression of cardiovascular disease (CVD) in older adults, with an emphasis on subclinical measures. The study recruited 5,888 adults aged 65 or older at entry in four U.S. communities and conducted extensive annual clinical exams between 1989-1999 along with semi-annual phone calls, events adjudication, and subsequent data analyses and publications (4). Additional data are collected by studies ancillary to CHS. In June 1990, four Field Centers (Sacramento, CA; Hagerstown, MD; Winston-Salem, NC; Pittsburgh, PA) completed the recruitment of 5201 participants. Between November 1992 and June 1993, an additional 687 participants, primarily African American, were recruited using similar methods. Blood samples were drawn from all participants at their baseline examination and during follow-up clinic visits, and DNA was subsequently extracted from available samples. CHS analyses were limited to participants with available DNA who consented to genetic studies. The baseline examinations consisted of a home interview and a clinic examination that assessed not only traditional risk factors but also measures of subclinical disease, including carotid ultrasound, echocardiography, electrocardiography, and pulmonary function. Between enrollment and 1998-99, participants were seen in the clinic annually and contacted by phone at 6-month intervals to collect information about hospitalizations and potential cardiovascular events.

Major exam components were repeated during annual follow-up examinations through 1999. Standard protocols for the identification and adjudication of events were implemented during follow-up. The adjudicated events are CHD, angina, heart failure (HF), stroke, transient ischemic attack (TIA), claudication and mortality. Adjudication of cause of death continues using a streamlined protocol; adjudication of other events ended in June 2015. Since 1999, participants have been contacted every 6 months by phone, primarily to ascertain health status and for events follow-up. The study was initially approved by institutional review boards at the Field Centers (Wake Forest, University of California – Davis, Johns Hopkins University, University of Pittsburgh), the Core Laboratory (University of Vermont) and at the Coordinating Center (University of Washington). The University of Washington now handles CHS Data Repository approvals.

#### Blood pressure measurements methods

Research staff who received central training in blood pressure measurement assessed repeat right-arm seated systolic and diastolic blood pressure levels at baseline with a Hawksley random-zero sphygmomanometer. Means of the repeated blood pressure measurements from the baseline examination were used for these analyses. Blood pressure lowering medication use was recorded from the medication history.

#### Ethics statement

All CHS participants provided informed consent, and the study was approved by the Institutional Review Board [or ethics review committee] of University Washington.

#### CHS acknowledgements

Cardiovascular Health Study: This research was supported by contracts HHSN268201200036C, HHSN268200800007C, HHSN268201800001C, N01HC55222, N01HC85079, N01HC85080, N01HC85081, N01HC85082, N01HC85083, N01HC85086, 75N92021D00006, and grants U01HL080295 and U01HL130114 from the National Heart, Lung, and Blood Institute (NHLBI), with additional contribution from the National Institute of Neurological Disorders and Stroke (NINDS). Additional support was provided by R01AG023629 from the National Institute on Aging (NIA). A full list of principal CHS investigators and institutions can be found at CHS-NHLBI.org. The content is solely the responsibility of the authors and does not necessarily represent the official views of the National Institutes of Health.

### CARDIA

The Coronary Artery Risk Development in Young Adults study (dbGaP accession phs000285) is a prospective multicenter study with 5,115 adults Caucasian and African American participants of the age group 18-30 years at baseline, recruited from four centers at the baseline examination in 1985-1986 (5). The recruitment was done from the total community in Birmingham, AL, from selected census tracts in Chicago, IL and Minneapolis, MN; and from the Kaiser Permanente health plan membership in Oakland, CA. Nine examinations have been completed in the years 0, 2, 5, 7, 10, 15, 20, 25 and 30, with high retention rates (91%, 86%, 81%, 79%, 74%, 72%, 72%, and 71%, respectively) and written informed consent was obtained in each visit.

#### Blood pressure measurements methods

Seated BP was measured on the right arm following 5 minutes rest using a random-zero sphygmomanometer. SBP and DBP were recorded as Phase I and Phase V Korotkoff sounds. Three measurements were taken at 1 minute intervals with the average of the second and third measurements taken for the BP values.

#### Ethics statement

All CARDIA participants provided informed consent, and the study was approved by the Institutional Review Boards of the University of Alabama at Birmingham and the University of Texas Health Science Center at Houston.

#### CARDIA acknowledgements

The Coronary Artery Risk Development in Young Adults Study (CARDIA) is conducted and supported by the National Heart, Lung, and Blood Institute (NHLBI) in collaboration with the University of Alabama at Birmingham (HHSN268201800005I & HHSN268201800007I), Northwestern University (HHSN268201800003I), University of Minnesota (HHSN268201800006I), and Kaiser Foundation Research Institute (HHSN268201800004I). CARDIA was also partially supported by the Intramural Research Program of the National Institute on Aging (NIA) and an intra-agency agreement between NIA and NHLBI (AG0005).

### FHS

The Framingham Heart Study (dbGaP accession phs000007) began in 1948 with the recruitment of an original cohort of 5,209 men and women (mean age 44 years; 55 percent women). In 1971 a second generation of study participants was enrolled; this cohort (mean age 37 years; 52% women) consisted of 5,124 children and spouses of children of the original cohort. A third-generation cohort of 4,095 children of offspring cohort participants (mean age 40 years; 53 percent women) was enrolled in 2002-2005 and are seen every 4 to 8 years. Details of study designs for the three cohorts are summarized elsewhere (6–8). At each clinic visit, a medical history was obtained, and participants underwent a physical examination. Only study participants consented for genetic and non-genetic data are included. FHS has been approved by the Boston University IRB

#### Blood pressure measurements methods

Systolic and diastolic blood pressures were measured twice by a physician on the left arm of the resting and seated participants using a mercury column sphygmomanometer. The blood pressure measurements used in this study were obtained at examination one cycle for all three generation cohorts.

#### Ethics statement

The Framingham Heart Study was approved by the Institutional Review Board of the Boston University Medical Center. All study participants provided written informed consent.

#### FHS acknowledgements

The Framingham Heart Study (FHS) acknowledges the support of contracts NO1-HC-25195, HHSN268201500001I and 75N92019D00031 from the National Heart, Lung and Blood Institute and grant supplement R01 HL092577-06S1 for this research. We also acknowledge the dedication of the FHS study participants without whom this research would not be possible.

### GENOA

The Genetic Epidemiology Network of Arteriopathy (GENOA) study, a part of the Family Blood Pressure Program (FBPP Investigators, 2002), consists of hypertensive sibships that were recruited for linkage and association studies in order to identify genes that influence blood pressure and its target organ damage (Daniels, 2004). In the initial phase of the GENOA study (Phase I: 1996-2001), all members of sibships containing ≥ 2 individuals with essential hypertension clinically diagnosed before age 60 were invited to participate, including both hypertensive and normotensive siblings. In the second phase of the GENOA study (Phase II: 2000-2004), 1,239 non-Hispanic white and 1,482 African American participants were successfully re-recruited to measure potential target organ damage due to hypertension.

#### Blood pressure measurements methods

SBP and DBPs were measured using an automated oscillometric BP measurement device with a consistent protocol across the FBPP networks. BP was measured three times on each participant by trained and certified technicians and then averaged for use in this analysis.

#### Ethics statement

Written informed consent was obtained from all subjects and approval was granted by participating institutional review boards (University of Michigan, University of Mississippi Medical Center, and Mayo Clinic).

#### GENOA acknowledgements

Support for the Genetic Epidemiology Network of Arteriopathy (GENOA) was provided by the National Heart, Lung and Blood Institute (U01 HL054457, U01 HL054464, U01 HL054481, R01 HL119443, and R01 HL087660) of the National Institutes of Health. We would like to thank the GENOA participants.

### HCHS/SOL

The Hispanic Community Health Study/Study of Latinos (dbGaP accession phs000810) is a community-based longitudinal cohort study of 16,415 self-identified Hispanic/Latino persons aged 18–74 years and selected from households in predefined census-block groups across four US field centers (in Chicago, Miami, the Bronx, and San Diego). The census-block groups were chosen to provide diversity among cohort participants with regard to socioeconomic status and national origin or background (9, 10). The HCHS/SOL cohort includes participants who self-identified as having a Hispanic/Latino background; the largest groups are Central American (n = 1,730), Cuban (n = 2,348), Dominican (n = 1,460), Mexican (n = 6,471), Puerto Rican (n = 2,728), and South American (n = 1,068). The HCHS/SOL baseline clinical examination occurred between 2008 and 2011 and included comprehensive biological, behavioral, and sociodemographic assessments. Visit 2 took place between 2014 and 2017, which re-examined 11,623 participants from the baseline sample. Visit 3 has started in 2020 and will last 3 years. In addition to clinic visit, participants are contacted annually to assess clinical outcomes. The study was approved by the Institutional Review Boards at each participating institution and written informed consent was obtained from all participants.

#### Blood pressure measurements methods

Blood pressure was measured on the right arm using an OMRON HEM-907 XL (Omron Healthcare, Inc., Lake Forest, IL) automatic sphygmomanometer, with the participant in the seated position and the arm resting. Cuff sizes were determined by measurement of the upper arm circumference. Four cuff sizes were available. Three blood pressures measurements were obtained 1 minute apart following an initial 5-minute rest period. The average of these 3 blood pressure values was used in this analysis. If there were fewer than 3 measurements, all available measurements were averaged. For more details, see (11).

#### Ethics statement

This study was approved by the institutional review boards (IRBs) at each field center, where all participants gave written informed consent, and by the Non-Biomedical IRB at the University of North Carolina at Chapel Hill, to the HCHS/SOL Data Coordinating Center. All IRBs approving the study are: Non-Biomedical IRB at the University of North Carolina at Chapel Hill. Chapel Hill, NC; Einstein IRB at the Albert Einstein College of Medicine of Yeshiva University. Bronx, NY; IRB at Office for the Protection of Research Subjects (OPRS), University of Illinois at Chicago. Chicago, IL; Human Subject Research Office, University of Miami. Miami, FL; Institutional Review Board of San Diego State University. San Diego, CA.

#### HCHS/SOL acknowledgements

The Hispanic Community Health Study/Study of Latinos is a collaborative study supported by contracts from the National Heart, Lung, and Blood Institute (NHLBI) to the University of North Carolina (HHSN268201300001I / N01-HC-65233), University of Miami (HHSN268201300004I / N01-HC-65234), Albert Einstein College of Medicine (HHSN268201300002I / N01-HC-65235), University of Illinois at Chicago – HHSN268201300003I / N01-HC-65236 Northwestern Univ), and San Diego State University (HHSN268201300005I / N01-HC-65237). The following Institutes/Centers/Offices have contributed to the HCHS/SOL through a transfer of funds to the NHLBI: National Institute on Minority Health and Health Disparities, National Institute on Deafness and Other Communication Disorders, National Institute of Dental and Craniofacial Research, National Institute of Diabetes and Digestive and Kidney Diseases, National Institute of Neurological Disorders and Stroke, NIH Institution-Office of Dietary Supplements.

### JHS

The Jackson Heart Study (<u>dbGaP accession phs000286)</u> is a longitudinal investigation of genetic and environmental risk factors associated with the disproportionate burden of cardiovascular disease in African Americans (12, 13). JHS is funded by the NHLBI and the National Institute on Minority Health and Health Disparities (NIMHD). JHS is an expansion of the ARIC study in its Jackson Field Center. At baseline, the JHS recruited 5306 African American residents of the Jackson Mississippi Metropolitan Statistical Area aged, approximately 6.6% of all African American adults aged 35-84 residing in the area. Participants were recruited via random sampling (17% of participants), volunteers (30%), prior participants in the Atherosclerosis Risk in Communities (ARIC) study (31%), and secondary family members (22%). Among these participants, approximately 3400 gave consent that allows genetic research. JHS participants received three back-to-back clinical examinations (Exam 1, 2000-2004; Exam 2, 2005-2008; and Exam 3, 2009-2013), and a fourth clinical examination has started in 2020. Participants are also contacted annually by telephone to update personal and health information including vital status, interim medical events, hospitalizations, functional status and sociocultural information

#### Blood pressure measurements methods

Two seated blood pressure measurements were taken using a Hawksley random zero sphygmomanometer and an appropriately sized cuff. BP measurements were calibrated using robust regression to the Omron HEM-907XL device (14). The use of antihypertensive medications was recorded in the medication history.

#### Ethics statement

The JHS study was approved by Jackson State University, Tougaloo College, and the University of Mississippi Medical Center IRBs, and all participants provided written informed consent.

#### JHS acknowledgements

The Jackson Heart Study (JHS) is supported and conducted in collaboration with Jackson State University (HHSN268201800013I), Tougaloo College (HHSN268201800014I), the Mississippi State Department of Health (HHSN268201800015I) and the University of Mississippi Medical Center (HHSN268201800010I, HHSN268201800011I and HHSN268201800012I) contracts from the National Heart, Lung, and Blood Institute (NHLBI) and the National Institute for Minority Health and Health Disparities (NIMHD). The authors also wish to thank the staffs and participants of the JHS.

### WHI

The Women’s Health Initiative (WHI) cohort. The WHI is a prospective national health study focused on identifying optimal strategies for preventing chronic diseases that are the major causes of death and disability in postmenopausal women [refs]. The WHI initially recruited 161,808 women between 1993 and 1997 with the goal of including a socio-demographically diverse population with racial/ethnic minority groups proportionate to the total minority population of US women aged 50-79 years. The WHI consists of two major parts: a set of randomized Clinical Trials and an Observational Study. The WHI Clinical Trials (CT; N=68,132) includes three overlapping components, each a randomized controlled comparison: the Hormone Therapy Trials (HT), Dietary Modification Trial, and Calcium and Vitamin D Trial. A parallel prospective observational study (OS; N = 93,676) examined biomarkers and risk factors associated with various chronic diseases. While the HT trials ended in the mid-2000s, active follow-up of the WHI-CT and WHI-OS cohorts has continued for over 25 years, with the accumulation of large numbers of diverse clinical outcomes, risk factor measurements, medication use, and many other types of data.

#### Blood pressure measurements methods

BP was measured by certified staff using standardized procedures and instruments(15). Two BP measures were recorded after 5 minutes rest using a mercury sphygmomanometer. Appropriate cuff bladder size was determined at each visit based on arm circumference. Diastolic BP was taken from the phase V Korotkoff measures. The average of the two measurements, obtained 30 seconds apart, was used in analyses

#### Ethics statement

All WHI participants provided informed consent and the study was approved by the Institutional Review Board (IRB) of the Fred Hutchinson Cancer Research Center.

#### WHI acknowledgements

The WHI program is funded by the National Heart, Lung, and Blood Institute, National Institutes of Health, U.S. Department of Health and Human Services through contracts 75N92021D00001, 75N92021D00002, 75N92021D00003, 75N92021D00004, 75N92021D00005.

### MESA

The Multi-Ethnic Study of Atherosclerosis (dbGaP accession phs000209) is a study of the characteristics of subclinical cardiovascular disease (disease detected non-invasively before it has produced clinical signs and symptoms) and the risk factors that predict progression to clinically overt cardiovascular disease or progression of the subclinical disease (16). MESA consisted of a diverse, population-based sample of an initial 6,814 asymptomatic men and women aged 45-84. 38 percent of the recruited participants were white, 28 percent African American, 22 percent Hispanic, and 12 percent Asian, predominantly of Chinese descent.

Participants were recruited from six field centers across the United States: Wake Forest University, Columbia University, Johns Hopkins University, University of Minnesota, Northwestern University and University of California - Los Angeles. Participants are being followed for identification and characterization of cardiovascular disease events, including acute myocardial infarction and other forms of coronary heart disease (CHD), stroke, and congestive heart failure; for cardiovascular disease interventions; and for mortality. The first examination took place over two years, from July 2000 - July 2002. It was followed by five examination periods that were 17-20 months in length. Participants have been contacted every 9 to 12 months throughout the study to assess clinical morbidity and mortality.

#### Blood pressure measurement methods

Resting BP was taken three times in the seated position after a five-minute rest using a Dinamap model Pro 100 automated oscillometric sphygmomanometer (Critikon, Tampa, Florida) (17) with the average of the last two measurements recorded and verified.

#### Ethics statements

All MESA participants provided written informed consent, and the study was approved by the Institutional Review Boards at The Lundquist Institute (formerly Los Angeles BioMedical Research Institute) at Harbor-UCLA Medical Center, University of Washington, Wake Forest School of Medicine, Northwestern University, University of Minnesota, Columbia University, and Johns Hopkins University.

#### MESA acknowledgements

MESA and the MESA SHARe project are conducted and supported by the National Heart, Lung, and Blood Institute (NHLBI) in collaboration with MESA investigators. Support for MESA is provided by contracts HHSN268201500003I, N01-HC-95159, N01-HC-95160, N01-HC-95161, N01-HC-95162, N01-HC-95163, N01-HC-95164, N01-HC-95165, N01-HC-95166, N01-HC-95167, N01-HC-95168, N01-HC-95169, UL1-TR-000040, UL1-TR-001079, UL1-TR-001420. MESA Family is conducted and supported by the National Heart, Lung, and Blood Institute (NHLBI) in collaboration with MESA investigators. Support is provided by grants and contracts R01HL071051, R01HL071205, R01HL071250, R01HL071251, R01HL071258, R01HL071259, and by the National Center for Research Resources, Grant UL1RR033176. The provision of genotyping data was supported in part by the National Center for Advancing Translational Sciences, CTSI grant UL1TR001881, and the National Institute of Diabetes and Digestive and Kidney Disease Diabetes Research Center (DRC) grant DK063491 to the Southern California Diabetes Endocrinology Research Center.

### MGB Biobank

Samples, genomic data, and health information were obtained from the Mass General Brigham (MGB) Biobank, a biorepository of consented patient samples at Mass General Brigham.

#### DNA samples

DNA samples are processed from whole blood that was collected as a dedicated research draw or as a clinical discard. Dedicated research samples are aimed to be processed within four hours of collection. Clinical discards are processed 24+ hours after collection. Whole blood is spun to buffy coat with a centrifuge and the buffy coat is stored in a freezer up to several months. The buffy coat is then extracted to DNA. The DNA is then placed in an ultralow freezer (-80°C).Each DNA aliquot contains a minimum of 2 ug of DNA. The concentration varies.

#### Genotyping

Samples have been genotyped using three versions of the biobank SNP array offered by Illumina that is designed to capture the diversity of genetic backgrounds across the globe. The first batch of data was generated on the Multi-Ethnic Genotyping Array (MEGA) array, the first release of this SNP array. The second, third, and fourth batches were generated on the Expanded Multi-Ethnic Genotyping Array (MEGA Ex) array. All remaining data were generated on the Multi-Ethnic Global (MEG) BeadChip.

#### Imputation

Prior to performing imputation, files were converted to VCF format, separated by chromosomes. When multiple probes measured the same genotypes, they were checked for concordance and were set to a missing value if the genotypes did not match. Files were uploaded to the Michigan Imputation Server, and Genotypes were imputed using TOPMed reference panel. Genomic coordinates are provided in GRCh38.

#### Quality control

We performed quality control using PLINK (v2.0. We filtered SNPs with low-quality imputation (r < 0.5), with missing call rates > 0.1, HWE p-value less than 1×10^-6^and MAF <1%.

We computed principal component (PC) using PLINK: we pruned the genotype data using a window size of 1000 variants, sliding across the genome with a step size of 250 variants at a time, filtering out any SNPs with LD R^2^>0.1. We used unrelated individuals (3rd degree, identified using PLINK) to compute the loadings for the first 10 PCs.

#### PRS construction

We constructed PRS using PRSice 2, using the same SNPs as those based on clumping performed on the TOPMed dataset, and otherwise the same methodology. Including, for comparability, when scaling PRS by mean and SD we used the ones estimated on the TOPMed dataset from stage 2.

#### Curated Disease Populations

We used outcomes from “curated disease populations”. These phenotypes were developed by the Biobank Portal team using both structured and unstructured electronic medical record (EMR) data and clinical, computational and statistical methods. Natural Language Processing (NLP) was used to extract data from narrative text. Chart reviews by disease experts helped identify features and variables associated with particular phenotypes and were also used to validate results of the algorithms. The process produced robust phenotype algorithms that were evaluated using metrics such as sensitivity, the proportion of true positives correctly identified as such, and positive predictive value (PPV), the proportion of individuals classified as cases by the algorithm (18). The high throughput phenotyping algorithm is as follows:

1. Create an initial phenotype definition using ICD-9 diagnosis codes.
2. Broaden the definition by determining the most up-to-date features (comorbidities, symptoms, medications) that create a more accurate profile of the phenotype when combined with ICD-9 codes. Features are extracted from online medical literature and knowledge bases via an Automated Feature Extraction Protocol (AFEP).
3. Narrow and refine the definition by determining the features that occur most often in the Biobank data. Extract, code, and rank features contained in clinical narratives with Natural Language Processing (NLP).
4. Create a gold-standard patient set for training the method. Query coded EMR data for the set of patients having at least one ICD-9 code for the phenotype. Apply a statistical sampling algorithm to select a random subset of those patients for full chart review. A clinical expert performs a full chart review to classify the patients as positive or negative for the phenotype.
5. Train a statistical model that incorporates all features in the definition to predict the presence or absence of the phenotype against the gold-standard patient set.
6. Apply the trained model to the entire Biobank Population.

The following table provides the prevalence and AUC of the phenotypes that we used (AUC was computed based on step 4 above).

| Phenotype | Biobank Prevalence | AUC |
| --- | --- | --- |
| Coronary Artery Disease | 13.5% | 0.989 |
| Hypertension | 42.0% | 0.912 |
| Ischemic Stroke | 3.1% | 0.907 |
| Type-II Diabetes Mellitus | 10.6% | 0.977 |
| Obesity | 48.9% | 0.948 |

#### Association analyses with disease outcomes

We used the outcomes above in association analyses. We adjusted for current age, sex, genotype batch, and the first 10 PCs. We only used unrelated individuals in the analysis and therefore used standard logistic regression models.

#### Ethics statement

All Biobank subjects have provided their consent to join the Partners Biobank, which includes agreeing to provide a blood sample linked to the electronic medical record. Subjects also agree to be recontacted by the Partners Biobank staff as needed.

#### Acknowledgements

We thank Mass General Brigham Biobank for providing samples, genomic data, and health information data.

## TOPMed and CCDG acknowledgements

Molecular data for the Trans-Omics in Precision Medicine (TOPMed) program was supported by the National Heart, Lung and Blood Institute (NHLBI). Genome sequencing for “NHLBI TOPMed - NHGRI CCDG: The BioMe Biobank at Mount Sinai” (phs001644) were performed at the Baylor College of Medicine Human Genome Sequencing Center and at the McDonnell Genome Institute (3UM1HG008853-WGS 01S2, HHSN26820160003 WGS 3I, HHSN26820160003 WGS 7I). Genome sequencing for “NHLBI TOPMed: Whole Genome Sequencing and Related Phenotypes in the Framingham Heart Study” (phs000974.v4.p3) was performed at the Broad Institute Genomics Platform (3R01HL092577-06S1, 3U54HG003067-12S2). DNA extraction for “NHLBI TOPMed: Genetic Epidemiology Network of Arteriopathy” (phs001345.v2.p1) was performed at the Mayo Clinic Genotyping Core and Genome sequencing was performed at the Broad Institute Genomics Platform (HHSN268201500014C) and the Northwest Genomics Center (3R01HL055673-18S1). Genome sequencing for “NHLBI TOPMed: The Jackson Heart Study” (phs000964.v1.p1) was performed at the Northwest Genomics Center (HHSN268201100037C). Genome sequencing for the “NHLBI TOPMed: The Atherosclerosis Risk in Communities Study” (phs001211.v3.p2) was performed at the Broad Institute Genomics Platform (3R01HL092577-06S1) and the Baylor College of Medicine Human Genome Sequencing Center (HHSN268201500015C, 3U54HG003273-12S2). Genomics sequencing for “NHLBI TOPMed: Cardiovascular Health Study” (phs001368.v2.p1) was performed at the Baylor College of Medicine Human Genome Sequencing Center (3U54HG003273-12S2, HHSN268201500015C, HHSN268201600033I). Genome sequencing for “NHLBI TOPMed: Hispanic Community Health Study/Study of Latinos” (phs001395.v1.p1) was performed at the Baylor College of Medicine Human Genome Sequencing Center (HHSN268201600033I). Genome sequencing for “NHLBI TOPMed: Women’s Health Initiative (WHI)” (phs001237.v2.p1) was performed at the Broad Institute of MIT and Harvard (HHSN268201500014C). Genome sequencing for “NHLBI TOPMed: Multi-Ethnic Study of Atherosclerosis” (phs001416.v2.p1) was performed at the Baylor College of Medicine Human Genome Sequencing Center (HHSN268201500015C and 3U54HG003273-12S2) and the Broad Institute for MIT and Harvard (3R01HL092577-06S1). Genome Sequencing for the NHLBI TOPMed: CARDIA Study (phs001612) was performed at the Baylor College of Medicine Human genome Sequencing Center (contract HHSN268201600033I). Core support including centralized genomic read mapping and genotype calling, along with variant quality metrics and filtering were provided by the TOPMed Informatics Research Center (3R01HL-117626-02S1; contract HHSN268201800002I). Core support including phenotype harmonization, data management, sample-identity QC, and general program coordination were provided by the TOPMed Data Coordinating Center (R01HL-120393; U01HL-120393; contract HHSN268201800001I). We gratefully acknowledge the studies and participants who provided biological samples and data for TOPMed. The Genome Sequencing Program (GSP) was funded by the National Human Genome Research Institute (NHGRI), the National Heart, Lung, and Blood Institute (NHLBI), and the National Eye Institute (NEI). The GSP Coordinating Center (U24 HG008956) contributed to cross-program scientific initiatives and provided logistical and general study coordination. The Centers for Common Disease Genomics (CCDG) program was supported by NHGRI and NHLBI, and whole genome sequencing was performed at the Baylor College of Medicine Human Genome Sequencing Center (UM1 HG008898 and R01HL059367).

## References

1. NCD Risk Factor Collaboration (NCD-RisC). Worldwide trends in blood pressure from 1975 to 2015: a pooled analysis of 1479 population-based measurement studies with 19·1 million participants. Lancet. 2017 Jan 7;389(10064):37–55.

2. Kearney PM, Whelton M, Reynolds K, Muntner P, Whelton PK, He J. Global burden of hypertension: analysis of worldwide data. Lancet. 2005 Jan 21;365(9455):217–23.

3. Graham I, Atar D, Borch-Johnsen K, Boysen G, Burell G, Cifkova R, et al. European guidelines on cardiovascular disease prevention in clinical practice: executive summary. Fourth Joint Task Force of the European Society of Cardiology and other societies on cardiovascular disease prevention in clinical practice (constituted by representatives of nine societies and by invited experts). Eur J Cardiovasc Prev Rehabil. 2007 Sep;14 Suppl 2:E1–40.

4. Lim SS, Vos T, Flaxman AD, Danaei G, Shibuya K, Adair-Rohani H, et al. A comparative risk assessment of burden of disease and injury attributable to 67 risk factors and risk factor clusters in 21 regions, 1990-2010: a systematic analysis for the Global Burden of Disease Study 2010. Lancet. 2012 Dec 15;380(9859):2224–60.

5. Shimbo D, Newman JD, Schwartz JE. Masked hypertension and prehypertension: diagnostic overlap and interrelationships with left ventricular mass: the Masked Hypertension Study. Am J Hypertens. 2012 Jun;25(6):664–71.

6. Pickering TG. The effects of environmental and lifestyle factors on blood pressure and the intermediary role of the sympathetic nervous system. J Hum Hypertens. 1997 Aug;11 Suppl 1:S9–18.

7. Bazzano LA, Green T, Harrison TN, Reynolds K. Dietary approaches to prevent hypertension. Curr Hypertens Rep. 2013 Dec;15(6):694–702.

8. Mills KT, Bundy JD, Kelly TN, Reed JE, Kearney PM, Reynolds K, et al. Global Disparities of Hypertension Prevalence and Control: A Systematic Analysis of Population-Based Studies From 90 Countries. Circulation. 2016 Aug 9;134(6):441–50.

9. Evangelou E, Warren HR, Mosen-Ansorena D, Mifsud B, Pazoki R, Gao H, et al. Genetic analysis of over 1 million people identifies 535 new loci associated with blood pressure traits. Nat Genet. 2018 Oct;50(10):1412–25.

10. Giri A, Hellwege JN, Keaton JM, Park J, Qiu C, Warren HR, et al. Trans-ethnic association study of blood pressure determinants in over 750,000 individuals. Nat Genet. 2019 Jan;51(1):51–62.

11. International Consortium for Blood Pressure Genome-Wide Association Studies, Ehret GB, Munroe PB, Rice KM, Bochud M, Johnson AD, et al. Genetic variants in novel pathways influence blood pressure and cardiovascular disease risk. Nature. 2011 Oct 6;478(7367):103–9.

12. Warren HR, Evangelou E, Cabrera CP, Gao H, Ren M, Mifsud B, et al. Genome-wide association analysis identifies novel blood pressure loci and offers biological insights into cardiovascular risk. Nat Genet. 2017 Mar;49(3):403–15.

13. Levy D, Ehret GB, Rice K, Verwoert GC, Launer LJ, Dehghan A, et al. Genome-wide association study of blood pressure and hypertension. Nat Genet. 2009 Jun;41(6):677–87.

14. Liu C, Kraja AT, Smith JA, Brody JA, Franceschini N, Bis JC, et al. Meta-analysis identifies common and rare variants influencing blood pressure and overlapping with metabolic trait loci. Nat Genet. 2016 Sep 12;48(10):1162–70.

15. Sofer T, Baier LJ, Browning SR, Thornton TA, Talavera GA, Wassertheil-Smoller S, et al. Admixture mapping in the Hispanic Community Health Study/Study of Latinos reveals regions of genetic associations with blood pressure traits. PLoS One. 2017 Nov 20;12(11):e0188400.

16. Liang J, Le TH, Edwards DRV, Tayo BO, Gaulton KJ, Smith JA, et al. Single-trait and multi-trait genome-wide association analyses identify novel loci for blood pressure in African-ancestry populations. PLoS Genet. 2017 May 12;13(5):e1006728.

17. Franceschini N, Fox E, Zhang Z, Edwards TL. Genome-wide association analysis of blood-pressure traits in African-ancestry individuals reveals common associated genes in African and non-African …. The American Journal of. 2013;

18. Liu Z, Shriner D, Hansen NF, Rotimi CN, Mullikin JC, NISC Comparative Sequencing Program. Admixture mapping identifies genetic regions associated with blood pressure phenotypes in African Americans. PLoS One. 2020 Apr 21;15(4):e0232048.

19. Kato N, Takeuchi F, Tabara Y, Kelly TN, Go MJ, Sim X, et al. Meta-analysis of genome-wide association studies identifies common variants associated with blood pressure variation in east Asians. Nat Genet. 2011 Jun;43(6):531–8.

20. Sofer T, Wong Q, Hartwig FP, Taylor K, Warren HR, Evangelou E, et al. Genome-Wide Association Study of Blood Pressure Traits by Hispanic/Latino Background: the Hispanic Community Health Study/Study of Latinos. Sci Rep. 2017 Sep 4;7(1):10348.

21. Choi SW, Mak TS-H, O’Reilly PF. Tutorial: a guide to performing polygenic risk score analyses. Nat Protoc. 2020 Sep;15(9):2759–72.

22. Choi SW, O’Reilly PF. PRSice-2: Polygenic Risk Score software for biobank-scale data. Gigascience. 2019 Jul 1;8(7).

23. Martin AR, Kanai M, Kamatani Y, Okada Y, Neale BM, Daly MJ. Clinical use of current polygenic risk scores may exacerbate health disparities. Nat Genet. 2019 Mar 29;51(4):584–91.

24. Duncan L, Shen H, Gelaye B, Meijsen J, Ressler K, Feldman M, et al. Analysis of polygenic risk score usage and performance in diverse human populations. Nat Commun. 2019 Jul 25;10(1):3328.

25. Grinde KE, Qi Q, Thornton TA, Liu S, Shadyab AH, Chan KHK, et al. Generalizing polygenic risk scores from Europeans to Hispanics/Latinos. Genet Epidemiol. 2019;43(1):50–62.

26. Vaura F, Kauko A, Suvila K, Havulinna AS, Mars N, Salomaa V, et al. Polygenic risk scores predict hypertension onset and cardiovascular risk. Hypertension. 2021 Apr;77(4):1119– 27.

27. Kanai M, Akiyama M, Takahashi A, Matoba N, Momozawa Y, Ikeda M, et al. Genetic analysis of quantitative traits in the Japanese population links cell types to complex human diseases. Nat Genet. 2018 Mar;50(3):390–400.

28. Cavazos TB, Witte JS. Inclusion of variants discovered from diverse populations improves polygenic risk score transferability. HGG Adv. 2021 Jan 14;2(1).

29. Taliun D, Harris DN, Kessler MD, Carlson J, Szpiech ZA, Torres R, et al. Sequencing of 53,831 diverse genomes from the NHLBI TOPMed Program. Nature. 2021;590(7845):290– 9.

30. Stilp AM, Emery LS, Broome JG, Buth EJ, Khan AT, Laurie CA, et al. A System for Phenotype Harmonization in the NHLBI Trans-Omics for Precision Medicine (TOPMed) Program. Am J Epidemiol. 2021 Apr 16;

31. Whelton PK, Carey RM, Aronow WS, Casey DE, Collins KJ, Dennison Himmelfarb C, et al. 2017 acc/aha/aapa/abc/acpm/ags/apha/ash/aspc/nma/pcna guideline for the prevention, detection, evaluation, and management of high blood pressure in adults: A report of the american college of cardiology/american heart association task force on clinical practice guidelines. Hypertension. 2018 Jun;71(6).

32. Conomos MP, Reiner AP, Weir BS, Thornton TA. Model-free Estimation of Recent Genetic Relatedness. Am J Hum Genet. 2016 Jan 7;98(1):127–48.

33. Hirata M, Kamatani Y, Nagai A, Kiyohara Y, Ninomiya T, Tamakoshi A, et al. Cross-sectional analysis of BioBank Japan clinical data: A large cohort of 200,000 patients with 47 common diseases. J Epidemiol. 2017 Mar;27(3S):S9–21.

34. Willer CJ, Li Y, Abecasis GR. METAL: fast and efficient meta-analysis of genomewide association scans. Bioinformatics. 2010 Sep 1;26(17):2190–1.

35. Lawrence M, Gentleman R, Carey V. rtracklayer: an R package for interfacing with genome browsers. Bioinformatics. 2009 Jul 15;25(14):1841–2.

36. Gogarten SM, Sofer T, Chen H, Yu C, Brody JA, Thornton TA, et al. Genetic association testing using the GENESIS R/Bioconductor package. Bioinformatics. 2019 Dec 15;35(24):5346–8.

37. Sofer T, Zheng X, Laurie CA, Gogarten SM, Brody JA, Conomos MP, et al. Population Stratification at the Phenotypic Variance level and Implication for the Analysis of Whole Genome Sequencing Data from Multiple Studies. BioRxiv. 2020 Mar 5;

38. Sofer T, Shaffer JR, Graff M, Qi Q, Stilp AM, Gogarten SM, et al. Meta-Analysis of Genome-Wide Association Studies with Correlated Individuals: Application to the Hispanic Community Health Study/Study of Latinos (HCHS/SOL). Genet Epidemiol. 2016 Jun 3;40(6):492–501.

39. Robin X, Turck N, Hainard A, Tiberti N, Lisacek F, Sanchez J-C, et al. pROC: an open-source package for R and S+ to analyze and compare ROC curves. BMC Bioinformatics. 2011 Mar 17;12:77.

40. Tryka KA, Hao L, Sturcke A, Jin Y, Wang ZY, Ziyabari L, et al. NCBI’s Database of Genotypes and Phenotypes: dbGaP. Nucleic Acids Res. 2014 Jan;42(Database issue):D975–9.

41. Yu S, Liao KP, Shaw SY, Gainer VS, Churchill SE, Szolovits P, et al. Toward high-throughput phenotyping: unbiased automated feature extraction and selection from knowledge sources. J Am Med Inform Assoc. 2015 Sep;22(5):993–1000.

42. Lambert SA, Abraham G, Inouye M. Towards clinical utility of polygenic risk scores. Hum Mol Genet. 2019 Nov 21;28(R2):R133–42.

43. Chatterjee N, Shi J, García M. Developing and evaluating polygenic risk prediction models for stratified disease prevention. Closas.

44. Sofer T, Kurniansyah N, Granot-Hershkovitz E, Goodman MO, Tarraf W, Broce I, et al. Polygenic Risk Scores for Alzheimer’s Disease and Mild Cognitive Impairment in Hispanics/Latinos in the U.S: The Study of Latinos - Investigation of Neurocognitive Aging. medRxiv. 2021 Jan 9;

45. Albiñana C, Grove J, McGrath JJ, Agerbo E, Wray NR, Bulik CM, et al. Leveraging both individual-level genetic data and GWAS summary statistics increases polygenic prediction. Am J Hum Genet. 2021 Apr 30;

46. Privé F, Arbel J, Vilhjálmsson BJ. LDpred2: better, faster, stronger. Bioinformatics. 2020 Dec 16;

47. Mak TSH, Porsch RM, Choi SW, Zhou X, Sham PC. Polygenic scores via penalized regression on summary statistics. Genet Epidemiol. 2017 Sep;41(6):469–80.

48. Lip S, Padmanabhan S. Genomics of blood pressure and hypertension: extending the mosaic theory toward stratification. Can J Cardiol. 2020 May;36(5):694–705.

## References

1. Kanai M, Akiyama M, Takahashi A, Matoba N, Momozawa Y, Ikeda M, et al. Genetic analysis of quantitative traits in the Japanese population links cell types to complex human diseases. Nat Genet. 2018 Feb 5;50(3):390–400.

2. Evangelou E, Warren HR, Mosen-Ansorena D, Mifsud B, Pazoki R, Gao H, et al. Genetic analysis of over 1 million people identifies 535 new loci associated with blood pressure traits. Nat Genet. 2018 Sep 17;50(10):1412–25.

3. The ARIC Investigators. The Atherosclerosis Risk in Communities (ARIC) Study: design and objectives. . Am J Epidemiol. 1989 Apr;129(4):687–702.

4. Fried LP, Borhani NO, Enright P, Furberg CD, Gardin JM, Kronmal RA, et al. The Cardiovascular Health Study: design and rationale. Ann Epidemiol. 1991 Feb;1(3):263–76.

5. Friedman GD, Cutter GR, Donahue RP, Hughes GH, Hulley SB, Jacobs DR, et al. CARDIA: study design, recruitment, and some characteristics of the examined subjects. J Clin Epidemiol. 1988;41(11):1105–16.

6. Dawber TR, Kannel WB, Lyell LP. An approach to longitudinal studies in a community: the Framingham Study. Ann N Y Acad Sci. 1963 May 22;107:539–56.

7. Splansky GL, Corey D, Yang Q, Atwood LD, Cupples LA, Benjamin EJ, et al. The Third Generation Cohort of the National Heart, Lung, and Blood Institute’s Framingham Heart Study: design, recruitment, and initial examination. Am J Epidemiol. 2007 Jun 1;165(11):1328–35.

8. Kannel WB, Feinleib M, McNamara PM, Garrison RJ, Castelli WP. An investigation of coronary heart disease in families. The Framingham offspring study. Am J Epidemiol. 1979 Sep;110(3):281–90.

9. Lavange LM, Kalsbeek WD, Sorlie PD, Avilés-Santa LM, Kaplan RC, Barnhart J, et al. Sample design and cohort selection in the Hispanic Community Health Study/Study of Latinos. Ann Epidemiol. 2010 Aug;20(8):642–9.

10. Sorlie PD, Avilés-Santa LM, Wassertheil-Smoller S, Kaplan RC, Daviglus ML, Giachello AL, et al. Design and implementation of the Hispanic Community Health Study/Study of Latinos. Ann Epidemiol. 2010 Aug;20(8):629–41.

11. Sorlie PD, Allison MA, Avilés-Santa ML, Cai J, Daviglus ML, Howard AG, et al. Prevalence of hypertension, awareness, treatment, and control in the Hispanic Community Health Study/Study of Latinos. Am J Hypertens. 2014 Jun;27(6):793–800.

12. Wyatt SB, Diekelmann N, Henderson F, Andrew ME, Billingsley G, Felder SH, et al. A community-driven model of research participation: the Jackson Heart Study Participant Recruitment and Retention Study. Ethn Dis. 2003;13(4):438–55.

13. Taylor HA, Wilson JG, Jones DW, Sarpong DF, Srinivasan A, Garrison RJ, et al. Toward resolution of cardiovascular health disparities in African Americans: design and methods of the Jackson Heart Study. Ethn Dis. 2005;15(4 Suppl 6):S6–4.

14. Seals SR, Colantonio LD, Tingle JV, Shimbo D, Correa A, Griswold ME, et al. Calibration of blood pressure measurements in the Jackson Heart Study. Blood Press Monit. 2019 Apr 16;

15. Design of the Women’s Health Initiative clinical trial and observational study. The Women’s Health Initiative Study Group. Control Clin Trials. 1998 Feb;19(1):61–109.

16. Bild DE, Bluemke DA, Burke GL, Detrano R, Diez Roux AV, Folsom AR, et al. Multi-Ethnic Study of Atherosclerosis: objectives and design. Am J Epidemiol. 2002 Nov 1;156(9):871– 81.

17. Ramsey M. Blood pressure monitoring: automated oscillometric devices. J Clin Monit. 1991 Jan;7(1):56–67.

18. Yu S, Liao KP, Shaw SY, Gainer VS, Churchill SE, Szolovits P, et al. Toward high-throughput phenotyping: unbiased automated feature extraction and selection from knowledge sources. J Am Med Inform Assoc. 2015 Sep;22(5):993–1000.

